# Fecal microbiota transplantation promotes immunotherapy sensitivity in refractory gastrointestinal cancer patients: open label, single-arm, single center, phase 1 study

**DOI:** 10.1101/2024.08.21.24312340

**Authors:** Yifan Zhang, Xiaomin Xu, Shulin Wang, Xiaochen Yin, Bohan Zhang, Zhengnong Zhu, Rujie Ji, Jing Zhu, Hermione He, Siyuan Cheng, Zihan Han, Tong Xie, Xiaotian Zhang, Yakun Wang, Si Shen, Yan Kou, Siyu Bao, Yingyu Liu, Baoran Cao, Christophe Bonny, Eran Segal, Yan Tan, Lin Shen, Zhi Peng

**Affiliations:** State Key Laboratory of Holistic Integrative Management of Gastrointestinal Cancers, Beijing Key Laboratory of Carcinogenesis and Translational Research, Department of Gastrointestinal Oncology, Peking University Cancer Hospital & Institute, Beijing 100142, China; Key Laboratory of Carcinogenesis and Translational Research (Ministry of Education), Department of Gastrointestinal Oncology, Peking University Cancer Hospital & Institute, Beijing 100142, China; Shenzhen Xbiome Biotech Co.Ltd, Shenzhen 518055, China; Department of Medical Oncology and Radiation Sickness, Peking University Third Hospital, Beijing 100191, China; Department of Colorectal Surgery, China-Japan Friendship Hospital, Beijing 100029, China; Department of Computer Science and Applied Mathematics, Weizmann Institute of Science, Rehovot, 7639302, Israel

**Keywords:** immnunotherapy resistance, fecal microbiota transplantation, advanced gastrointestinal cancer, colonization

## Abstract

**Background:** The discovery and therapeutic application of immune checkpoint inhibitors (ICIs) has significantly improved clinical outcomes in cancer treatment. However, the response rate is still low in gastrointestinal (GI) cancers. The gut microbiome’s impact on immune modulation is a promising area for enhancing ICI efficacy.

**Methods:** This study (NCT04130763) is an open label, single-arm, single center, phase 1 study assessing the safety and efficacy of fecal microbiota transplantation (FMT) from healthy donors in ten advanced GI cancer patients resistant to anti-PD-(L)1 treatment. Patients received initial FMT treatment via oral capsules, followed by a combination therapy phase, where maintenance FMT was paired with nivolumab at 3mg/kg every two weeks for six cycles. Serial biomarker assessments were conducted through both fecal and blood sampling.

**Findings:** The combination of FMT and anti-PD1 treatment was well tolerated with no serious adverse reactions observed among all 10 patients. The objective response rate was 20% and the disease control rate was 40%. The progression-free survival of these two responders were 15 and more than 19 months respectively. Clinical benefits were associated with colonization of donor-derived immunogenic microbes, and an activated immune status reflected by peripheral immune cell populations. Responder-enriched microbes interacted closely as a butyrate-functional guild, while non-responder-enriched microbes interacted sparsely and had higher fraction of oral-originated microbes. Donor-specific microbial traits that influence clinical efficacy of FMT were validated in an independent cohort.

**Interpretation:** The current study demonstrates the feasibility of FMT for ICI-refractory GI cancer patients and provides a foundation for live biotherapeutic product (LBP) development to enhance ICI efficacy.

## Introduction

GLOBOCAN database^1^ indicates 19.3 million new cancers cases and about 9.96 million deaths from cancers worldwide in 2020. Gastrointestinal (GI) cancers, including esophageal, gastric, and colorectal cancers, accounted for nearly 19% of cancer incidence and more than 22% of mortality. Notably, Asia leads in GI cancer cases, accounting for 80%, 75% and 52% new cases of esophageal, gastric and colorectal cancer, respectively.^1^ Immune checkpoint inhibitors (ICIs) have succeeded as a new treatment option for patients with GI cancer where commonly used anti-cancer treatments, such as surgery, radiation, and chemotherapy, have historically shown unsatisfactory efficacy.^2^ However, the response rate of ICI monotherapy is 11%−15% for GI cancers (phase III trial results),^3,4^ particularly lower in gastric and esophageal cancer patients with low PD-L1 expression, as well as gastric and colorectal cancer patients with microsatellite stable status. Moreover, a high percentage (42%-71%) of cancer patients initially responsive to ICIs eventually develop resistance,^5^ substantially limiting the ICI efficacy in terms of overall survival (OS) or progression-free survival (PFS).^6–8^ This represents a large unmet medical need and highlights the urgency for novel therapeutic strategies to overcome immunotherapy resistance.

Previous studies^9–13^ have demonstrated the association between gut microbiome and ICI efficacy, with evidence that microbiome from ICI responders or healthy donors could enhance the efficacy of anti-PD-1 therapy through microbe-derived small molecules such as short chain fatty acids (SCFA), inosine, arginine and neoantigen mimicry etc.^10,14,15^ Germ-free mice given fecal transplants from immunotherapy responders have shown improved tumor control and T cell responses.^11^Moreover, multiple research groups have demonstrated fecal microbiota transplantation (FMT) from ICI-responsive patients can overcome resistance in metastatic melanoma, with an objective response rate (ORR) of 20-30%.^12,13^ Recently, safety and efficacy have been demonstrated from administration of healthy donor’s FMT in combination with ICI therapy on anti-PD-1 naive melanoma patients,^5^ achieving an ORR of 65%.

While FMT has demonstrated promising results in clinical studies for “hot tumors” with high mutational burdens that respond rapidly to ICI, such as melanoma, its role in “cold tumors”, such as GI cancers requires further investigation. Therefore, we initiated an open-label, single-arm, single center, Phase 1 trial to investigate the safety and efficacy of ICI treatment combined with FMT derived from healthy donors as a combination therapy for GI cancer patients resistant to ICI.

## Methods

### Study Design and Patient Population

This study (Clinical trial registration number: NCT04130763) is an open label, single-arm, single center, phase 1 study that included patients with advanced GI cancer who were resistant or refractory to ICI treatment. This study was approved by the Ethics Committee at Peking University Cancer Hospital. The clinical protocol is available online. GI patients with an Eastern Cooperative Oncology Group (ECOG) performance status of 0-1, and consent to serial biomarker assessments via blood and fecal sampling were included in the study. Ten patients were enrolled and treated at a single center in Beijing, China.

### Study Treatment

Patients received initial FMT treatment via oral capsules (per capsule: 0.7 g fecal extract). Total 60 capsules were administered within the first week of the study, followed by a combination therapy phase, where maintenance FMT was paired with nivolumab at 3mg/kg every two weeks for six cycles (Fig.1a). For those showing positive response, the combination treatment was extended with full details described in Supplementary Material.

**Figure 1:**
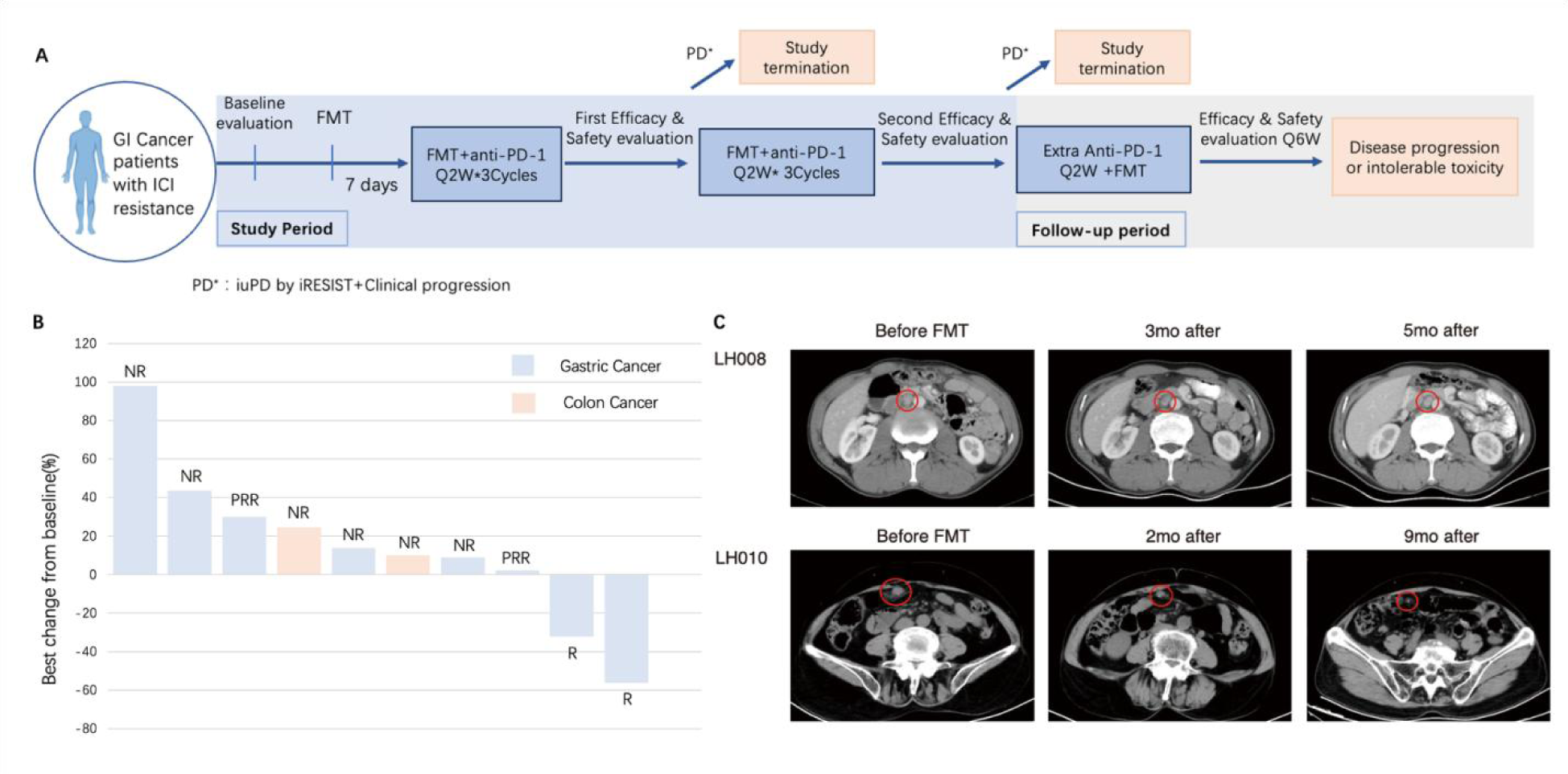
Study design and clinical outcomes. **a.** Flow chart depicting the clinical trial protocol. Healthy donors were screened with general donor qualification criteria (Supplementary Material). Study participants were given an initial FMT treatment one week prior beginning combination therapy with FMT (maintenance FMT) and anti-PD-1 therapy (nivolumab). **b.** Waterfall plot showing the response as the change in size of target lesions from baseline over time from FMT and anti-PD-1 therapy according to iRECIST criteria. **c.** Representative CT scans from the two Responders.GI: gastrointestinal; FMT: fecal microbiota transplantation; iRECIST: immune Response Evaluation Criteria in Solid Tumors; ICI: immune checkpoint inhibitor.

### Clinical assessment

FMT and anti-PD-1 related toxicities were evaluated using Common Terminology Criteria for Adverse Events (CTCAE) 5.0 grading scale. iRECIST was used as the main efficacy evaluation criteria for all imaging evaluations. Per protocol, responder (R) is defined as patients without icPD(immune confirmed progressive disease) or clinical progression per efficacy evaluation at any evaluation point. Partial responder (PRR) is defined as patients with iSD(immune stable disease) or better efficacy outcome at the first efficacy evaluation (after 6 weeks of treatment) but has icPD(immune confirmed progressive disease) or clinical progression within 4-8 weeks thereafter. Non-responder (NR) is defined as patients who had icPD or iuPD(immune unconfirmed progressive disease) with clinical progression at any evaluation point.

### Gut microbiome analysis

Sequenced metagenomic reads were quality controlled using Fastp^16^. The low-quality portion of reads were trimmed and reads longer than 50 bp were retained. Host DNA was removed using KneadData^17^. Functional profiling was performed using HUMAnN 3.0^18^ against MetaCyC database. GutSMASH^19^ was applied for primary metabolic pathway prediction. Alpha-diversity was calculated using Shannon index, Observed, and inverse Simpson index and Beta-diversity was measured with Bray-curtis distance.Strain-level analysis was conducted with StrainPhIAN3^18^. Based on the state of strains at baseline and after treatment, we defined the strain level colonization concepts of “strain gain” and “strain replace” (sup fig 1c). A genomics-based strain analysis method^20^ was also applied to confirm strain level colonization (Supplementary Material).

### Statistical analysis

Permutational multivariate ANOVA (PERMANOVA) was used to identify microbiome composition differences between groups (p<0.05). Differential species between Rs and NRs were identified using LEfSe^21^ (LDA > 2 and p< 0.05) and MaAsLin2^22^ (adjusted.p <0.3), while LogFold Change (|logFold| > 2) was measured in comparing donors. Wilcoxon signed rank test was performed for SCFA productivity comparison(p<0.05). Linear mixed effect model^23^ (adjust.p <0.3) was applied for group comparison with multiple samples from the same subject. Weighted gene co-expression network analysis (WGCNA)^24,25^ was employed for co-occurrence analysis and the network was visualized by Cytoscape.^26^

For TCR data, Pearson correlation was executed to measure the association between frequency of top 100 CDR3aa and TCR CDR3aa Shannon index (p<0.05). Multi-omics analysis was performed with samples collected from baseline to mid-term evaluation using Spearman correlation (adjusted.p <0.3).

Two 10-fold cross validation XGBoost classifiers were generated in an independent validation cohort. Average AUC of 10 models was used to represent the classifier performance.

All statistical analysis and plotting were performed using R v3.6.3 and machine learning was performed using python 3.9.

## Results

### Clinical Outcomes: Efficacy

The average interval from the last dose of previous ICI treatment to the first dose of anti-PD-1 monoclonal antibody (nivolumab) + FMT combination therapy was 64·9 days, ensuring the observed responses were attributable to the new regimen rather than a delayed effect of previous treatments. Patient baseline characteristics were summarized in Table 1. The ORR was 20% (Fig. 1b) and the disease control rate (DCR) was 40%. Notably, all patients who responded to the combination therapy had MSS gastric cancer with low levels of PD-L1 expression. Therefore, when the analysis was restricted to gastric cancer patients, the ORR was 25% with DCR 50%. Two patients (LH008 and LH010) exhibited immune partial response (iPR) associated with distinct healthy FMT donors (donor 4 and 5, respectively). Patient LH008 achieved the best response of iPR with a PFS of 15 months. Patient LH010 sustained the iPR status for over 19 months treatment (Fig. 1c). The FMT capsule administration frequency for LH010 was adjusted from every ICI cycle to every four cycles, with ongoing monitoring for treatment efficacy and survival. These findings indicate the anti-PD-1+FMT combination therapy could enhance the responsiveness in GI cancer patients, who were previously unresponsive to anti-PD-(L)1 therapies.

**Table 1.** Patient baseline Characteristics.

| **Table 1 Patient baseline Characteristics** |  |  |  |  |  |
| --- | --- | --- | --- | --- | --- |
| **Baseline Characteristics** | **R (n=2)** | **PRR (n=2)** | **R+PRR (n=4)** | **NR (n=6)** | **Total** |
| **Age, years** |  |  |  |  |  |
| ≤65 n(%) | 1(50·0) | 0 | 1(25·0) | 4(66·7) | 5 (50·0) |
| >65 n(%) | 1(50·0) | 2(100·0) | 3(75·0) | 2(33·3) | 5 (50·0) |
| **Gender** |  |  |  |  |  |
| Male n(%) | 1(50·0) | 1(50·0) | 2(50·0) | 6(100·0) | 8 (80·0) |
| Female n(%) | 1(50·0) | 1(50·0) | 2(50·0) | 0 | 2 (20·0) |
| **Disease Duration (m)** |  |  |  |  |  |
| Mean (SD) | 26·79 (18·09) | 34·69 (32·61) | 30·74 (22·01) | 14·28 (7·42) | 20·86 (16·26) |
| Medium (Q1,Q3) | 26·79(13·99,39·58) | 34·69(11·63, 57·75) | 26·79(12·81, 48·66) | 11·60(8·31, 22·01) | 13·62(9·95, 24·87) |
| **Tumor Diagnosis** |  |  |  |  |  |
| GC n(%) | 2(100·0) | 2(100·0) | 4(100·0) | 4(66·7) | 8(80·0) |
| CRC n(%) | 0 | 0 | 0 | 2(33·3) | 2 (20·0) |
| **PD-L1 Expression (CPS)** |  |  |  |  |  |
| ≥10 | 0 | 0 | 0 | 2(33·3) | 2(20·0) |
| ≥1 | 2(100·0) | 1(50·0) | 3(75·0) | 6(100·0) | 9(90·0) |
| **Length of IO treatment (m)** |  |  |  |  |  |
| Mean (SD) | 9·35 (1·93) | 10·58 (0·79) | 9·96 (1·40) | 4·48 (2·09) | 6·67 (3·33) |
| Medium (Q1,Q3) | 9·35(7·98, 10·71) | 10·58(10·02, 11·14) | 10·36(9·00, 10·92) | 3·88(3·55, 4·17) | 6·08(3·68, 10·02) |
| **First Dose since Last dose of previous IO regimen, days** |  |  |  |  |  |
| Mean (SD) | 57·5 (21·92) | 37·5 (4·95) | 47·5 (17·37) | 76·5 (25·18) | 64·9 (26·02) |
| Medium (Q1,Q3) | 57·5(42·0, 73·0) | 37·5(34·0, 41·0) | 41·5(37·5, 57·5) | 72·5(69·0, 102·0) | 70·5(41·0, 73·0) |
| **Previous lines of regimen** |  |  |  |  |  |
| 1 line n(%) | 1(50·0) | 1(50·0) | 2(50·0) | 3(50·0) | 5 (50·0) |
| 2 lines n(%) | 0 | 1(50·0) | 1(25·0) | 1(16·7) | 2 (20·0) |
| 3 lines or more n(%) | 1(50·0) | 0 | 1(25·0) | 2 (33·3) | 3 (30·0) |
| **Previous IO regimen** |  |  |  |  |  |
| Monotherapy n(%) | 1(50·0) | 1(50·0) | 2(50·0) | 5(83·3) | 7(70·0) |
| Combination therapy n(%) | 1(50·0) | 1(50·0) | 2(50·0) | 1(16·7) | 3(30·0) |

R: Responder; PRR: Partial-Responder (Protocol defined); NR: Non responder; GC: gastric cancer; CRC: colorectal cancer; MSS: microsatellite stable; CPS: Combined Positive Score; IO: immune-oncology.

### Clinical outcomes: Safety

The combination therapy used in the current study was well tolerated with no serious adverse reactions during the study; no early termination from the study was due to adverse event (AE) (table S1-S3); and treatment-related AEs were minimal (table S4, S5). During the FMT monotherapy period, only one patient experienced treatment-related AEs (nausea and vomiting), which were mild and did not require treatment. During the combination therapy period, no AEs were reported by the investigator as being only related to FMT. Three patients experienced four mild treatment-related adverse events primarily affecting the gastrointestinal tract, including nausea and constipation. Overall, FMT combined with ICI was found to be safe and well tolerated in this cohort.

### Increased FMT colonization in responders

NRs had slightly higher alpha diversity compared to Rs and PRRs at baseline, but significance could not be assessed due to small sample size (Fig.2a). After FMT treatment, there was an increase in observed species across all groups (mean: pre = 120, post = 136, Fig.2a), matching the results of Routy’s et al.^5^ study, that species richness increased after FMT regardless of clinical outcome. Further considering the evenness of the whole bacteria community diversity, Rs showed higher increase than PRRs following FMT treatment, while NRs had a decreasing trend, indicating less rare species in Rs (Fig.2a).

**Figure 2:**
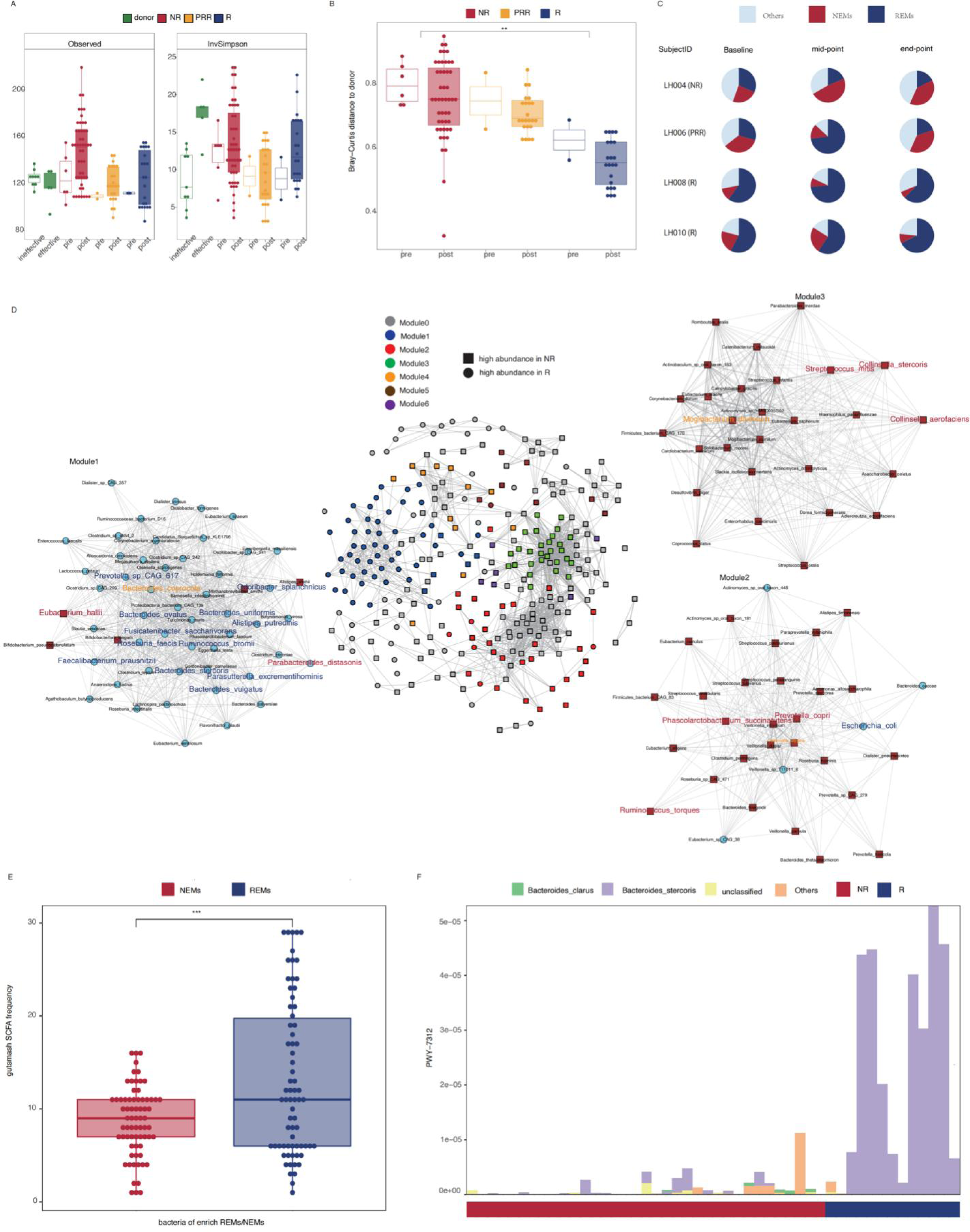
Gut microbiome changes associated with clinical outcome. **a.** Alpha diversity of patients pre and post FMT measured by Observed and InvSimpson. **b.**Bray-curtis distance to donor before and after the FMT treatment. **c.** REM and NEM composition through the treatment in three patients representing each clinical outcome group. **d.** WGCNA was used to generate a network of 293 species of bacteria using the 77 post-treatment samples from Rs and NRs. Bacteria with average higher abundance in Rs were presented as blue circle and those with average higher abundance in NRs were red triangles. The REMs and NEMs were presented in blue and red texts in each module respectively. The hubs of each module were highlighted in orange text. **e.** Comparison of SCFA production capacity between REMs and NEMs. **f.** *B. stercoris* associated PWY-7312 D-fucofuranose biosynthetic pathway was enriched in Rs post treatment. ns, not significant; *P < 0·05; **P < 0·01; ***P < 0·001

Beta diversity suggested significant differences between patients and healthy donors (p=0·001) but not between patients at baseline (p=0·141, sup fig.1a). After treatment, the gut microbiome composition of all patients was closer to their corresponding donors, with Rs showing significantly greater similarities compared to NRs (p=0·009) (Fig.2b).

### Specific colonized bacteria leading to a favorable response to ICI treatment in gastric cancer patients

The patients regained ICI sensitivity from FMT combination therapy were gastric cancer patients. Considering the potential difference in the mode of action between cancer types, we focused on gastric cancer patients to pinpoint FMT-induced microbial changes related to ICI efficacy(sup fig.2a).

Firstly, by considering both the microbial abundance and prevalence, the core microbiome^27^ of R and NR groups were identified. Secondly, samples collected at mid-term evaluation were stratified to detect response related differential bacteria. Thirdly, MaAsLin2^22^ was applied with samples collected along the treatment, while addressing the issue of data dependence. In total, we detected 14 species enriched in Rs (R-enriched-microbes, REMs) and 25 enriched in NRs (NR-enriched-microbes, NEMs) (sup fig.2a).

The ratio of REMs to NEMs serves as a balance indicator to assess the importance of these two microbial fractions. This indicator was significantly higher in Rs compared to NRs throughout FMT treatment (p = 0·008, Fig.3b, sup fig.2b), fluctuating greatly but generally matching the clinical outcomes. For patient LH006, the ratio initially increased but gradually decreased after two months, in line with the clinical outcome of an initial response followed by the development of resistance (Fig.3b).

**Figure 3:**
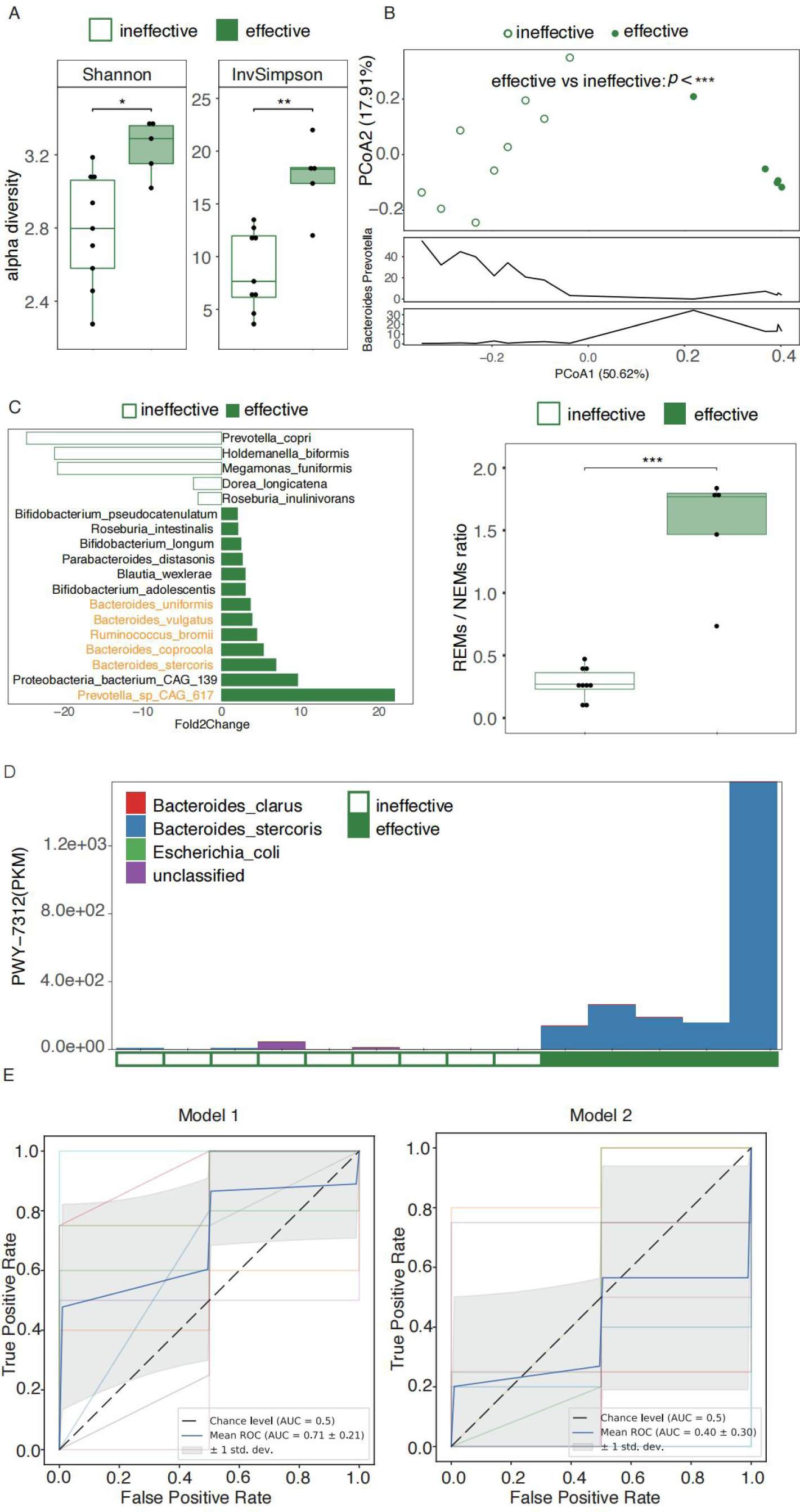
Validation of efficacy-related microbial traits in donors of this study (A-D) and an independent dataset (E). **a.** Comparison of alpha diversity at the species level between donors. **b.** Comparison of gut microbiome composition between donors by PERMANOVA. **c.** Differentially abundant bacteria between donors. Differential analysis was performed by comparing the foldchange (foldchange>2). 6 REM bacteria were enriched in effective donors and were highlighted in orange. **d.** B. stercoris associated PWY-7312 D-fucofuranose biosynthesis was higher in effective donors. **e**. Two 10-fold cross validation XGBoost classifiers was built to stratify gastric cancer patients by clinical outcome. AUC was calculated to measure the performance of the classifier. ns, not significant; *P < 0·05; **P < 0·01; ***P < 0·001

Of the 14 REMs, 7 species were not present at baseline, indicating the Rs might gain these species through FMT (sup fig.2c). At the strain level, total 10 REM strains were considered as engrafted bacteria (sup fig.2c). In terms of the 25 NEMs, 16 and 8 were considered engrafted bacteria at species and strain level respectively (sup fig.5).

Strain sharing of REMs between donors and recipients at the bacterial genomic level was also analyzed to trace the potential source of the REMs.^20^ A decrease in strain dissimilarity through treatment was observed (mean of strain dissimilarity: baseline= 0·009, post treatment = 0·002, sup fig.2d).

### Colonized bacteria formed a microbial network exerting immune-related functions

Microbes can interact with each other for specific biological functions that further impact clinical outcomes.^28^ Through network analysis,^24,25^ we observed REMs and NEMs were clearly separated as different modules in the whole microbiome network (Fig.2d). Bacteria from REMs formed a more intricate network than NEMs, with 13 REMs closely associated in module 1, whereas bacteria in NEMs dispersed across different modules. In module 1, *Bacteroides coprocola* served as the super-node, the one that connected the most to other group members. *Veillonella atypica* and *Mogibacterium diversum*, were the super-nodes in the two NEM represented modules (module 2 and 3), respectively (Fig.3c).

At the functional level, the potential capacity of short chain fatty acid (SCFA) production was significantly higher in REMs (p < 0·05, Fig.3d). We also observed two pathways enriched in Rs (table S6). The REM member *Bacteroides stercoris* was identified as the top contributor to pathway PWY-7312 D-fucofuranose biosynthesis (Fig. 3e).

### Donor-specific microbial traits influenced FMT clinical efficacy

To investigate donor-specific microbial characteristics associated with the clinical efficacy, the gut microbiome of the two Rs’ donors were compared to three NRs’ donors (see Table S7).

At the community level, Rs’ donors had higher alpha diversity (p < 0·05, Fig. 3a), and their gut microbiome compositions were significantly different from NRs’ donors (p = 0·001), mainly driven by an apparently lower ratio of *Prevotella* to *Bacteroides* (Fig.3b). We further identified 6 REMs had higher relative abundance in the Rs’ donors (foldchange >2, Fig. 3c). *B.stercoris* associated PWY-7312 D-fucofuranose biosynthesis was also higher in R’s donors (Fig. 3d). These results indicate that the enrichment of REMs and related functions, are potentially important factors forclinical outcomes.

### Large gastric cancer cohort validated key taxa and pathways associated with clinical efficacy

Validation was performed for the microbes and functional pathways identified in this study using an independent external dataset of gastrointestinal cancer patients.^14^ Sixty-three gastric cancer patients were included for validation, including 43 long-term responders and 20 primary resistance patients, corresponding respectively to Rs and NRs in our study (sup fig.3a).

Two XGBoost classifiers were built to verify whether REMs and NEMs comprise general microbial characteristics related to immunotherapy. The model using only the REMs and NEMs (mean AUC = 0·71) outperformed the model using all other bacteria (mean AUC = 0·4) (Fig.3e), indicating the composition of REMs and NEMs can accurately discriminate clinical responses in gastric cancer patients even without FMT intervention. In terms of functional level, *B.stercoris* remained as the main contributor of PWY-7312 D-fucofuranose biosynthetic pathway in long-term responders (sup fig.3b).

### Host immune profiles associated with the clinical response of FMT

Patient immune profiles revealed higher CDR3aa diversity in Rs and PRRs compared to NRs at baseline (Fig. 4a). CDR3aa quantity followed a similar pattern (sup fig.4a), with a strong correlation between TCR diversity and top 100 CDR3 sequence abundance (p<0·001, sup fig. 4b). Flow cytometry results indicated elevated CD4+ and CCR7+CD45RA-cell population along with the lowest FoxP3+ cell population in Rs. NRs had the lowest Ki-67+ cell percentageat baseline (Fig.4a), which persisted after combination treatment (table S8, S9). Notably, Rs showed reduced CD279+ cells after FMT (Fig. 4b).

**Figure 4:**
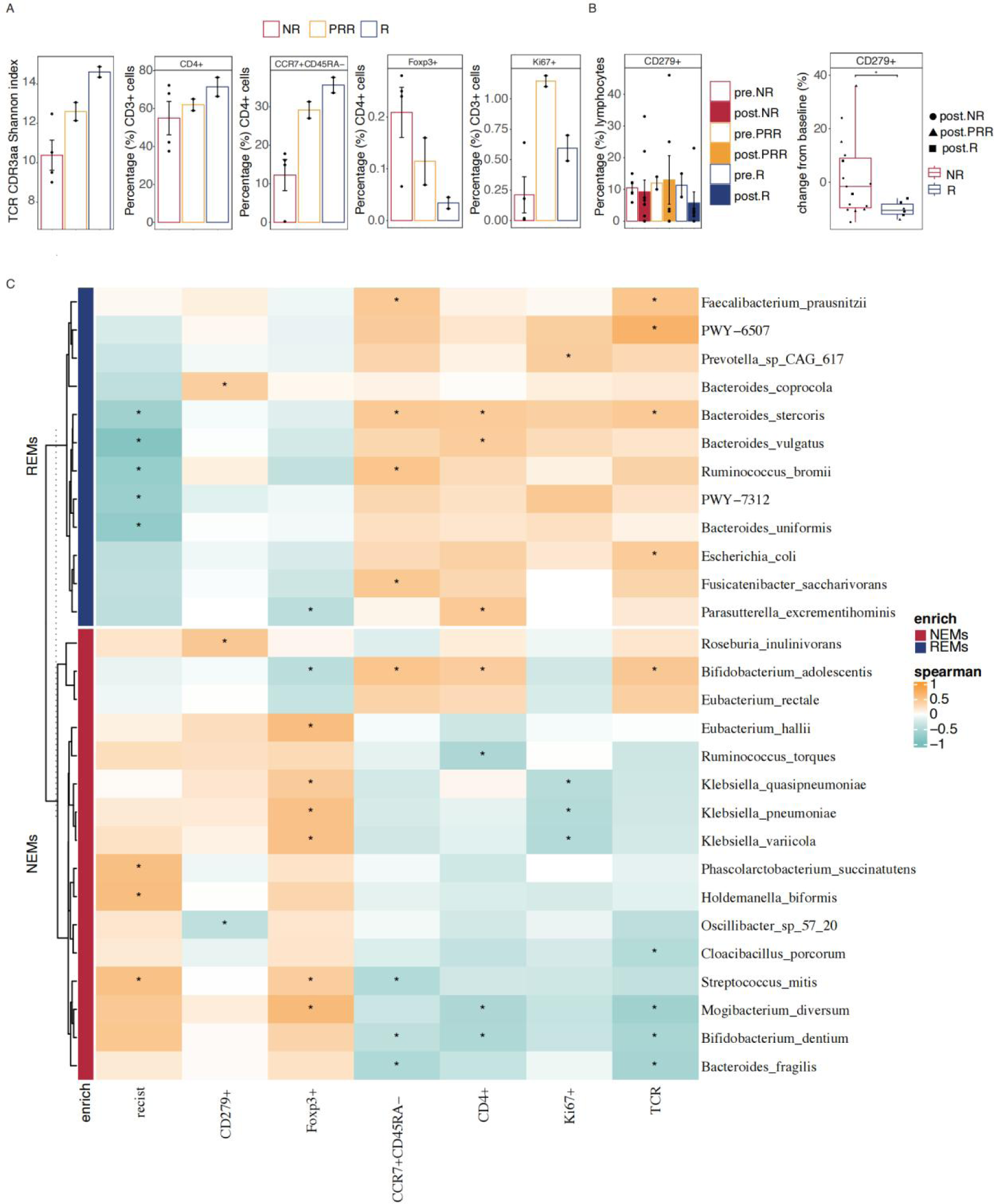
Host immune profiles associated with clinical responses. **a.** TCR diversity and immune cell sub-population of patients’ peripheral blood at baseline. Mean and SEM are shown. **b.** Comparison of CD279+ cells before and after PD-1+FMT therapy in NRs, PRRs and Rs analyzed by patients and samples repectively. **c.** Heatmap representing Spearman correlation between microbial features and host immune and tumor characteristics. Eight baseline samples and 16 post-treatment samples collected at the first clinical evaluation from each patient were used. REMs and NEMs were clustered on the left through complete-linkage. Color gradient of the cell indicates the correlation coefficient. Significant correlation is defined as an FDR adjusted p <0·3 and is labeled with “*” in the corresponding cell.

Spearman correlation was performed to investigate the potential relationship between immune responses and gut microbiome (Fig.4c). The relative abundance of REMs such as *B. stercoris* and *Faecalibacterium prausnitzii* had significantly positive correlations (adjust p.value < 0·3) with CCR7+CD45RA-cells, Ki-67 cells, CD4+ cells and TCR diversity. Among the NEMs such as *M. diversum* and *Streptococcus mitis*, the opposite pattern was observed. A significantly negative correlation was detected between Foxp3+ cells and the REM member *Parasutterella excrementihominis*. Among the NEMs, three species of *Klebsiella* showed significantly negative correlations with the quantity of Ki-67 cells but positively correlated with Foxp3+ cells.

## Discussion

Immune checkpoint inhibitors have significantly advanced cancer treatment in recent decades, yet their effectiveness in GI cancer has been unsatisfying. For instance, the ORR to Nivolumab in gastric cancer patients who had at least two lines of chemotherapy without prior ICI exposure was only 11.2%,^4^ with the median overall survival (mOS) of 5.26 months.^29^ Previous studies^30^ have predominantly attributed the diminished efficacy of immunotherapy to ICI resistance, underscoring an urgent need for innovative therapeutic strategies to surmount this resistance and reconfigure the treatment landscape for GI cancer patients.

The pivotal role of the gut microbiome in modulating host immunity has catalyzed investigations into its capacity to augment ICI efficacy, with pioneering studies employing FMT,^5,12,13^ bacterial consortia,^31,32^ live biotherapeutic products (LBPs), ^33^ or prebiotics.^34^ For the FMT applications, fecal samples from either responder patients or healthy donors were applied and exciting clinical outcomes were reported in “hot” melanoma population.^5,12,13^ Our study extends this inquiry to the realm of GI cancer patients who have become refractory to ICIs, a group often deemed “cold” to immunotherapy.^35^ The combination of FMT and anti-PD-1 therapy has demonstrated encouraging results in the current study (20% ORR for GI cancer, 25% gastric cancer),. When focusing on effective donors, the ORR escalates to 67%, and the one-year overall survival rate for gastric cancer patients reaches an impressive milestone of 100%, with no significant adverse events related to FMT. These results validate the treatment’s safety and effectiveness while revealing opportunities to delve deeper into the mechanisms of donor-recipient dynamics and the possibilities of tailored microbiome therapies.

The elevated immunotherapy sensitivity first attributed to the FMT engraftment. Our study revealed a significantly higher similarity of gut microbiome between responders and their respective donors post-treatment due to successful engraftment. Similar results have also been observed in previous FMT studies,^20,36^ supporting the fact that robust colonization is the first essential component in FMT success. Increased but not significant bacterial diversity after FMT was observed in our cohort in accordance with previous findings in FMT on melanoma and NSCLC patients.^37,38^ Aside from colonization rate, our results suggest rational donor selection is an important factor for FMT success as well, similar to Baruch et al’s finding in melanoma.^12^ In our study, effective donors harbored microbial signatures such as higher diversity, higher REM/NEM ratio, which could be further applied for rational donor selection.^37,38^ Co-occurrence analysis of FMT recipients indicated REMs and NEMs interacted closely within each respective group, forming several functional “guilds”^28^, which are microbial groups that exploit the same class of resources or work together as a coherent functional group.^28^ Most microbes in the “responsiveness” guild (module 1) were found to be strong SCFA producers, a physiologically important function related to host immunity.^39^ Several studies have shown butyrate treatment could modulate cytotoxic CD8+ T cell response, a major player in cancer treatment.^40–42^ We also observed an REM member’s associated signature (*B. stercoris*) related to fucofuranose biosynthesis. This sugar is usually found on bacterial surface and comprises the sugar moiety of Gilvocarcin V, a topoisomerase II inhibitor.^43–45^ Thus, the likelihood for bacterial derived fucofuranose contribution to the improvement of PD1 responsiveness is high and warrants further validation. Notably, we identified the enrichment of *B. stercoris* and fucofuranose biosynthesis pathway in effective donors of the current FMT intervention as well as long-term responders in another independent cohort, further supporting the robustness of this finding. NEMs, on the other hand, clustered into multiple modules. The super node of module 2 was *M.diversum,* an oral-orginated microbe^46^ and its abundance in the tongue coating microbiome was associated with increased risk of colorectal cancer^47^. Other oral-originated microbes (such as *S*. *mitis* and *Bifidobacterium dentium*) were also found in NEMs and significantly associated with host immune suppression status (Foxp3+, CCR7+CD45RA-, CD4+). Abnormal oral gut transmission has been reported to associate with a number of diseases, such as IBD, ^48^ colon cancer^49^ etc. Increased fraction of oral microbes in the NRs’ feces indicates a more dysbiosis gut microbiome ecosystem^50^ despite of FMT treatment, and this could also serve as a predictor for FMT efficacy.

Recipient immune status profiling revealed Rs exhibited a more activated immune environment compared with NRs, a trend that remained consistent even after 6 cycles of combination therapy. This suggests that more activated immune environment may create a favorable environment for FMT to exert its role in facilitating the anti-tumor effect of ICI. Further comparison between NR and R samples revealed a reduction of immune cells expressing PD-1 after combination treatment which suggests combination treatment increased PD-1 receptor occupancy (RO) by anti-PD-1 antibody and potential re-activated exhausted T cells. Significant increases of immune cell population in the circulatory system were not observed after combination treatment in Rs, indicating that immune activation primarily occurred in the tumor, with minimal impact on the peripheral circulatory profile. Consequently, the risk of excessive immune activation and potential immune-related disorders may be reduced. However, due to the lack of tumor tissue after combination treatment, the current study was unable to compare the immune microenvironment before and after FMT. Interestingly, a separate study corroborates our speculation by discovering a similar phenomenon that the impact of immunotherapy on cells circulating in peripheral blood was relatively minor compared to the notable alterations observed in tumors.^51^ Moreover, by integrating the host immune status and microbial traits, the current results suggest that effective donor FMT has the potential to modify microbiome composition and facilitate colonization of strains with immune activation properties in the recipient, thereby facilitating the anti-tumor effect of ICI.

The current study benchmarks the potential efficacy and excellent safety profile of FMT and highlights proper donor selection as an important tool toward clinical outcomes for ICI refractory GI cancer patients. Meanwhile, we acknowledge the limitation of the small cohort size, which impacts the statistical power but believe the application of stringent statistical methods would generate robust signals, and validation in an independent cohort of these signals are assuring. The microbial collection linked to ICI response may serve as a foundational basis for informed donor selection in future larger cohorts and holds substantial potential for rational microbial consortia development.

## Data Availability

All data produced in the present study are available upon reasonable request to the authors

## Acknowledgements

The authors gratefully thank all the patients and their families for participating in this study. The authors also thank Yong Liang for building the pipeline of metagenomics data analysis, Haopeng Lu for supporting the metagenomics sequencing of stool samples and Dr. Adam Arterbery for helping the English language editing of the manuscript.

## Data sharing

Data from this study are not publicly available. The study protocol hasbeen published, including details of sample handing and processing. Requests for access to the full dataset can be made by sending an email with a research plan to the corresponding author.

