## Supplementary material for "Fecal microbiota transplantation promotes immunotherapy sensitivity in refractory gastrointestinal cancer patients: open label, single-arm, single center, phase 1 study": sup material

#### Supplementary Materials Catalogue

**Sup Fig1**

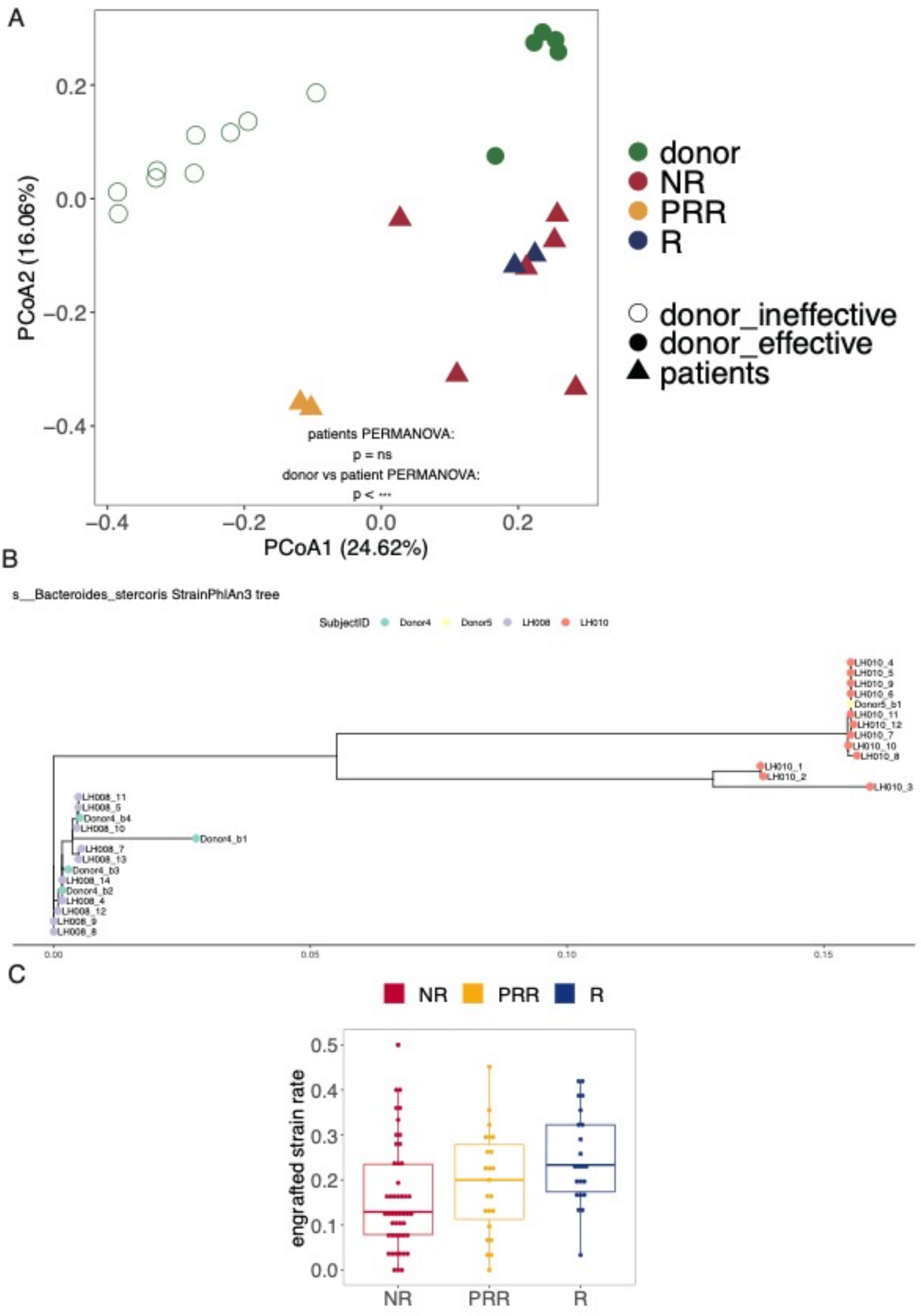

A
PCoA2 (16.06%)
PCoA1 (24.62%)
0.2
0.0
−0.2
−0.4
−0.4
−0.2
0.0
0.2
patients PERMANOVA:
p = ns
donor vs patient PERMANOVA:
p < ***
donor
NR
PRR
R
donor_ineffective
donor_effective
patients
B
s__Bacteroides_stercoris StrainPhlAn3 tree
SubjectID
Donor4
Donor5
LH008
LH010
LH010_4
LH010_5
LH010_9
LH010_6
Donor5_b1
LH010_11
LH010_12
LH010_7
LH010_10
LH010_8
LH010_1
LH010_2
LH010_3
LH008_11
LH008_5
Donor4_b4
LH008_10
Donor4_b1
LH008_7
LH008_13
Donor4_b3
LH008_14
Donor4_b2
LH008_4
LH008_12
LH008_9
LH008_8
0.00
0.05
0.10
0.15
C
NR
PRR
R
engrafted strain rate
0.5
0.4
0.3
0.2
0.1
0.0
NR
PRR
R

Sup Fig2

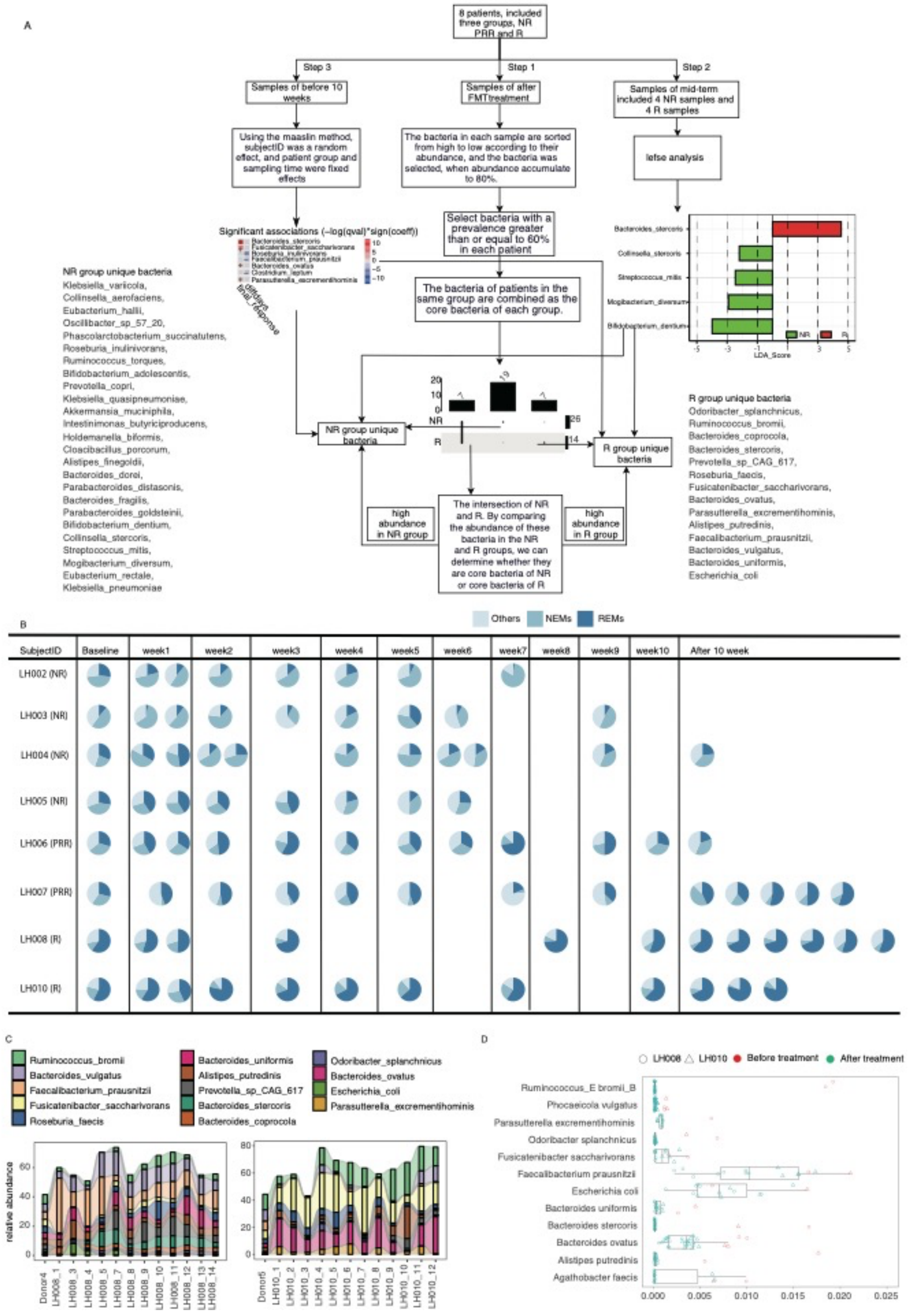
A
8 patients, included three groups, NR PRR and R
Step 3
Step 1
Step 2
Samples of before 10 weeks
Samples of after FMTtreatment
Samples of mid-term included 4 NR samples and 4 R samples
Using the maaslin method, subjectID was a random effect, and patient group and sampling time were fixed effects
The bacteria in each sample are sorted from high to low according to their abundance, and the bacteria was selected, when abundance accumulate to 80%.
lefse analysis
Significant associations (−log(qval)*sign(coeff))
Select bacteria with a prevalence greater than or equal to 60% in each patient
The bacteria of patients in the same group are combined as the core bacteria of each group.
NR group unique bacteria
R group unique bacteria
high abundance in NR group
The intersection of NR and R. By comparing the abundance of these bacteria in the NR and R groups, we can determine whether they are core bacteria of NR or core bacteria of R
high abundance in R group
NR group unique bacteria
Klebsiella_variicola, Collinsella_aerofaciens, Eubacterium_hallii, Oscillibacter_sp_57_20, Phascolarctobacterium_succinatutens, Roseburia_inulinivorans, Ruminococcus_torques, Bifidobacterium_adolescentis, Prevotella_copri, Klebsiella_quasipneumoniae, Akkermansia_muciniphila, Intestinimonas_butyriciproducens, Holdemanella_biformis, Cloacibacillus_porcorum, Alistipes_finegoldii, Bacteroides_dorei, Parabacteroides_distasonis, Bacteroides_fragilis, Parabacteroides_goldsteinii, Bifidobacterium_dentium, Collinsella_stercoris, Streptococcus_mitis, Mogibacterium_diversum, Eubacterium_rectale, Klebsiella_pneumoniae
R group unique bacteria
Odoribacter_splanchnicus, Ruminococcus_bromii, Bacteroides_coprocola, Bacteroides_stercoris, Prevotella_sp_CAG_617, Roseburia_faecis, Fusicatenibacter_saccharivorans, Bacteroides_ovatus, Parasutterella_excrementihominis, Alistipes_putredinis, Faecalibacterium_prausnitzii, Bacteroides_vulgatus, Bacteroides_uniformis, Escherichia_coli
LDA_Score
B
Others NEMs REMs
SubjectID Baseline week1 week2 week3 week4 week5 week6 week7 week8 week9 week10 After 10 week
LH002 (NR)
LH003 (NR)
LH004 (NR)
LH005 (NR)
LH006 (PRR)
LH007 (PRR)
LH008 (R)
LH010 (R)
C
Ruminococcus_bromii
Bacteroides_vulgatus
Faecalibacterium_prausnitzii
Fusicatenibacter_saccharivorans
Roseburia_faecis
Bacteroides_uniformis
Alistipes_putredinis
Prevotella_sp_CAG_617
Bacteroides_stercoris
Bacteroides_coprocola
Odoribacter_splanchnicus
Bacteroides_ovatus
Escherichia_coli
Parasutterella_excrementihominis
relative abundance
D
LH008 LH010 Before treatment After treatment
Ruminococcus_E bromii_B
Phocaeicola vulgatus
Parasutterella excrementihominis
Odoribacter splanchnicus
Fusicatenibacter saccharivorans
Faecalibacterium prausnitzii
Escherichia coli
Bacteroides uniformis
Bacteroides stercoris
Bacteroides ovatus
Alistipes putredinis
Agathobacter faecis

**Sup Fig3**

A

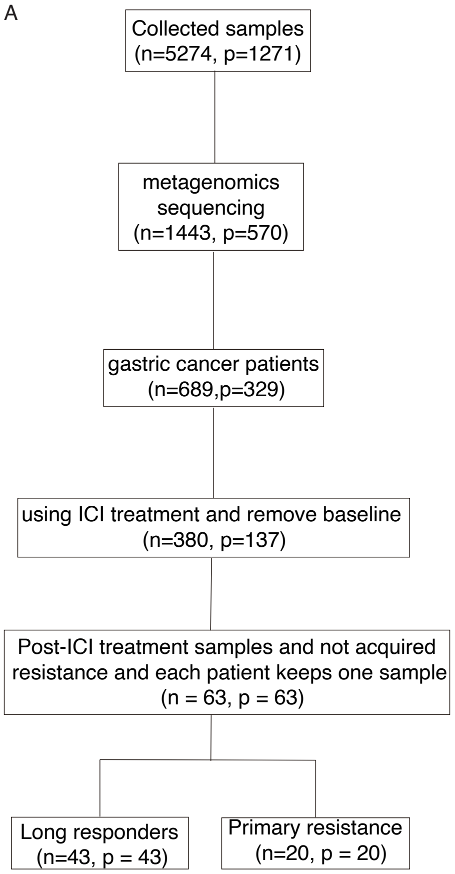
Collected samples
(n=5274, p=1271)
metagenomics sequencing
(n=1443, p=570)
gastric cancer patients
(n=689,p=329)
using ICI treatment and remove baseline
(n=380, p=137)
Post-ICI treatment samples and not acquired resistance and each patient keeps one sample
(n = 63, p = 63)
Long responders
(n=43, p = 43)
Primary resistance
(n=20, p = 20)

B

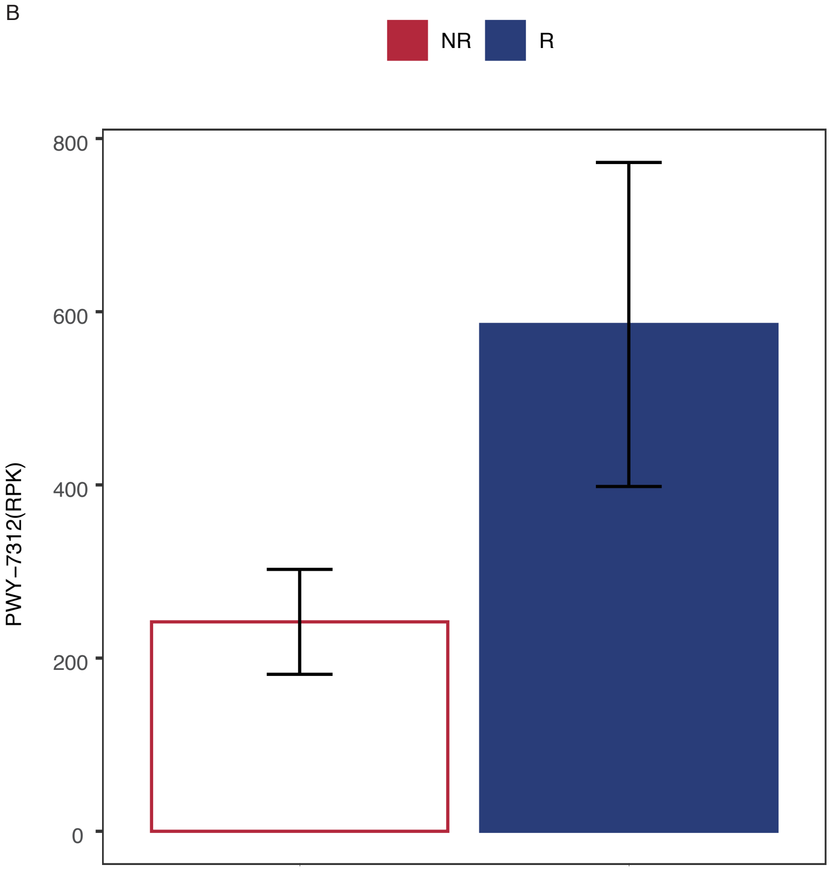
NR
R
PWY−7312(RPK)
800
600
400
200
0

**Sup Fig4**

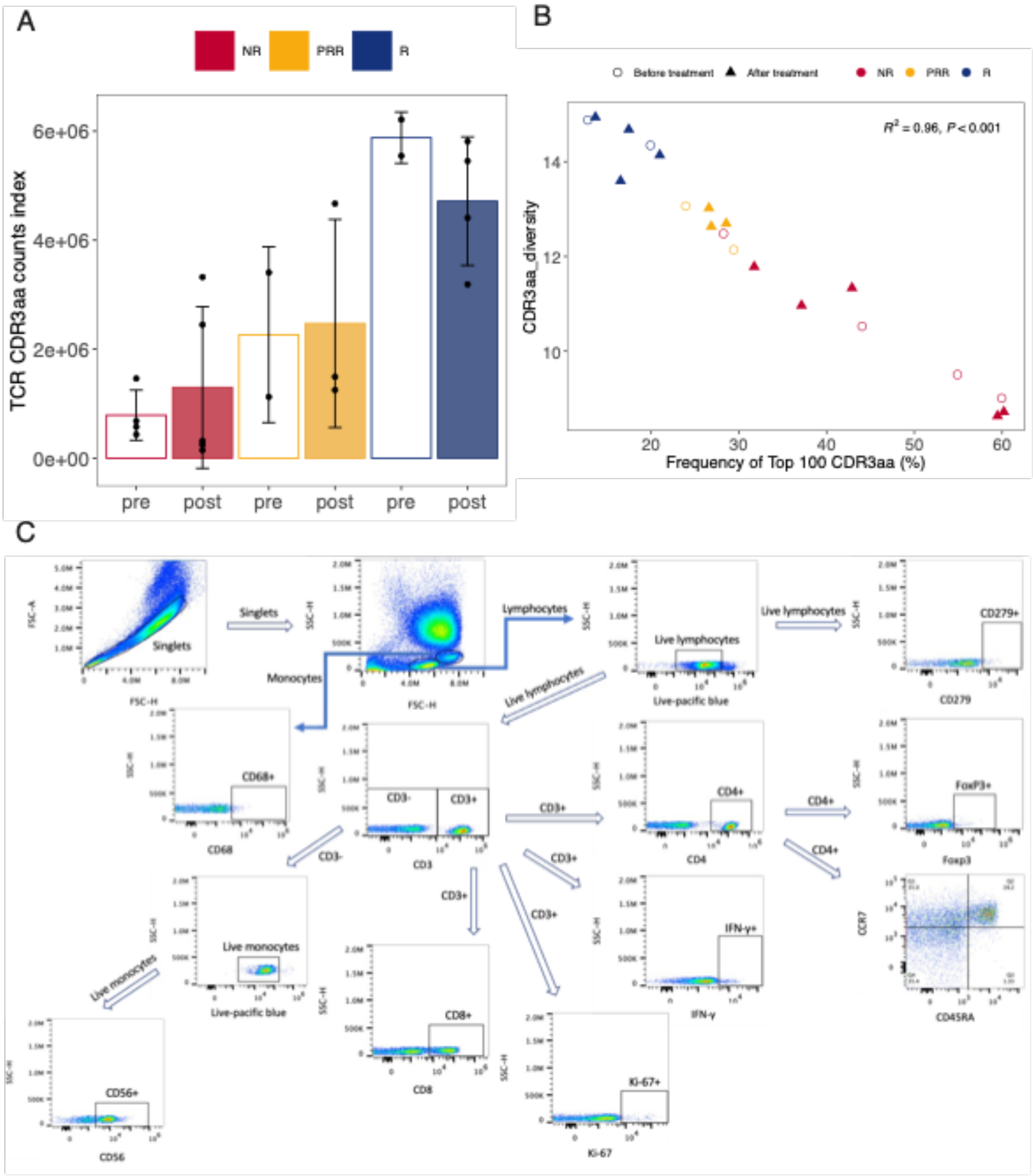

A
NR
PRR
R
TCR CDR3aa counts index
6e+06
4e+06
2e+06
0e+00
pre
post
pre
post
pre
post
B
Before treatment
After treatment
NR
PRR
R
$R^2 = 0.96,\ P < 0.001$
CDR3aa_diversity
14
12
10
20
30
40
50
60
Frequency of Top 100 CDR3aa (%)
C
Singlets
Lymphocytes
Monocytes
Live lymphocytes
Live-pacific blue
CD279+
CD279
CD68+
CD68
CD3-
CD3+
CD3
CD4+
CD4
FoxP3+
Foxp3
IFN-γ+
IFN-γ
CCR7
CD45RA
Live monocytes
CD8+
CD8
Ki-67+
Ki-67
CD56+
CD56

#### Sup Fig5a

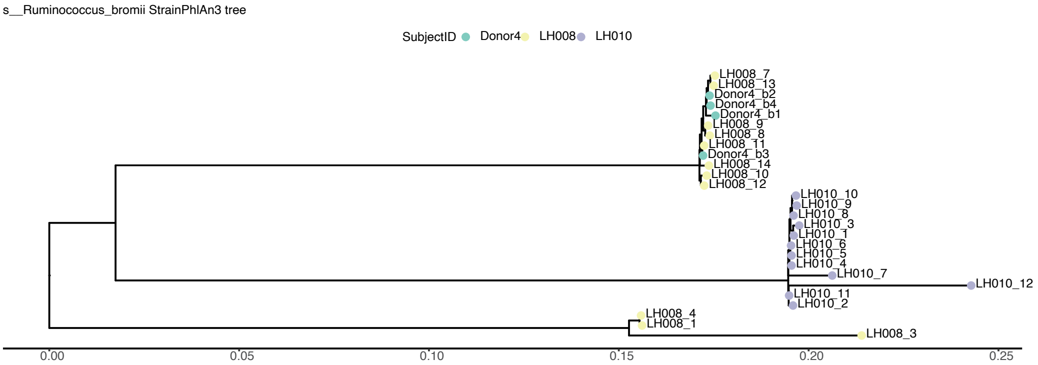
s__Ruminococcus_bromii StrainPhlAn3 tree
SubjectID Donor4 LH008 LH010
LH008_7
LH008_13
Donor4_b2
Donor4_b4
Donor4_b1
LH008_9
LH008_8
LH008_11
Donor4_b3
LH008_14
LH008_10
LH008_12
LH010_10
LH010_9
LH010_8
LH010_3
LH010_1
LH010_6
LH010_5
LH010_4
LH010_7
LH010_12
LH010_11
LH010_2
LH008_4
LH008_1
LH008_3
0.00
0.05
0.10
0.15
0.20
0.25

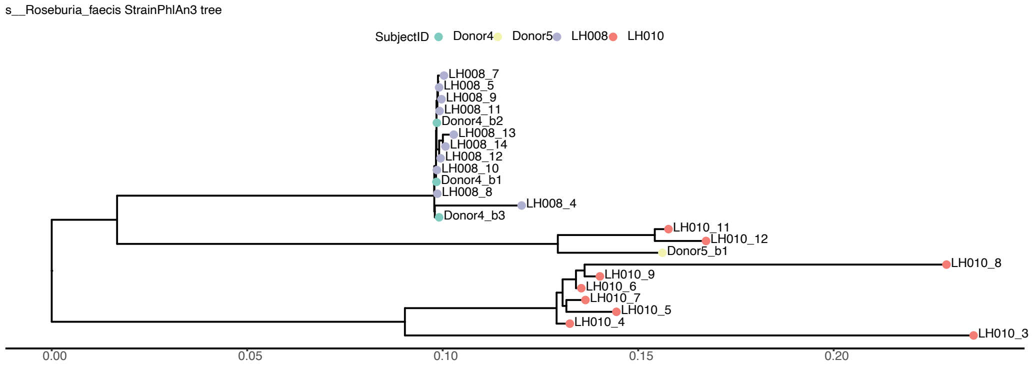
s__Roseburia_faecis StrainPhlAn3 tree
SubjectID Donor4 Donor5 LH008 LH010
LH008_7
LH008_5
LH008_9
LH008_11
Donor4_b2
LH008_13
LH008_14
LH008_12
LH008_10
Donor4_b1
LH008_8
LH008_4
Donor4_b3
LH010_11
LH010_12
Donor5_b1
LH010_8
LH010_9
LH010_6
LH010_7
LH010_5
LH010_4
LH010_3
0.00
0.05
0.10
0.15
0.20

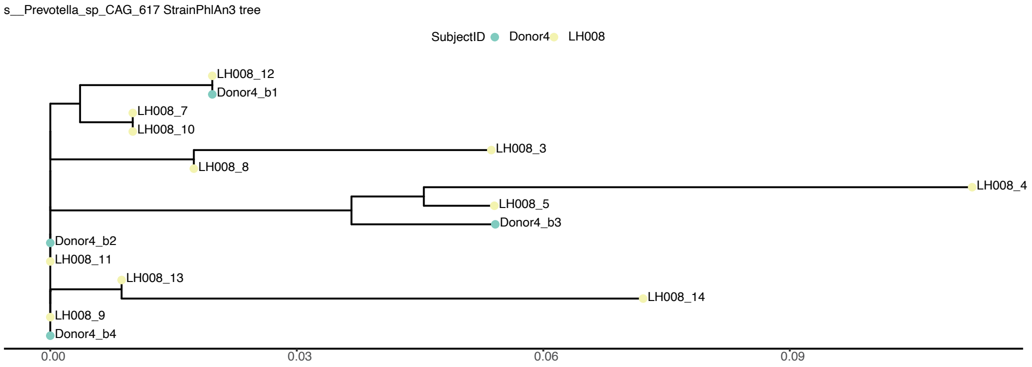
s__Prevotella_sp_CAG_617 StrainPhlAn3 tree
SubjectID Donor4 LH008
LH008_12
Donor4_b1
LH008_7
LH008_10
LH008_3
LH008_8
LH008_4
LH008_5
Donor4_b3
Donor4_b2
LH008_11
LH008_13
LH008_14
LH008_9
Donor4_b4
0.00
0.03
0.06
0.09

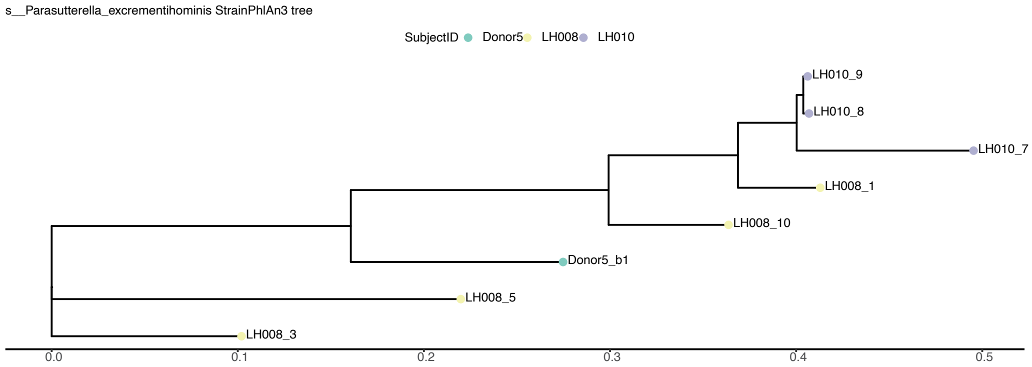
s__Parasutterella_excrementihominis StrainPhlAn3 tree
SubjectID Donor5 LH008 LH010
LH010_9
LH010_8
LH010_7
LH008_1
LH008_10
Donor5_b1
LH008_5
LH008_3
0.0
0.1
0.2
0.3
0.4
0.5

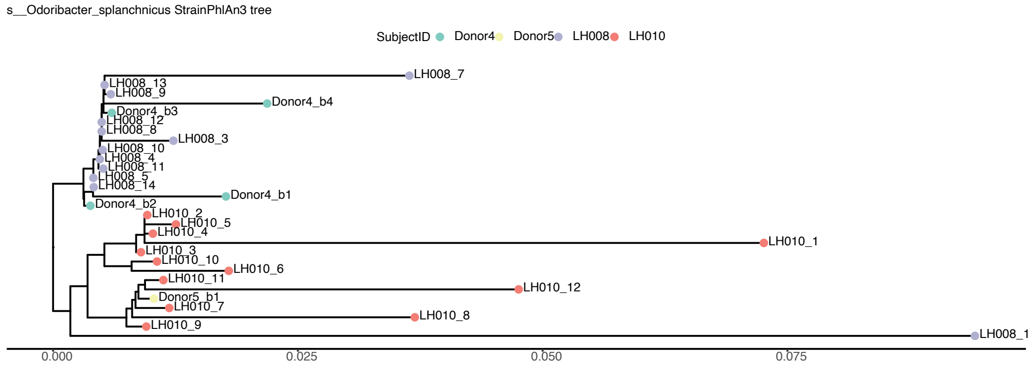
s__Odoribacter_splanchnicus StrainPhlAn3 tree
SubjectID Donor4 Donor5 LH008 LH010
LH008_7
LH008_13
LH008_9
Donor4_b4
Donor4_b3
LH008_12
LH008_8
LH008_3
LH008_10
LH008_4
LH008_11
LH008_5
LH008_14
Donor4_b1
Donor4_b2
LH010_2
LH010_5
LH010_4
LH010_1
LH010_3
LH010_10
LH010_6
LH010_11
LH010_12
Donor5_b1
LH010_7
LH010_8
LH010_9
LH008_1
0.000
0.025
0.050
0.075

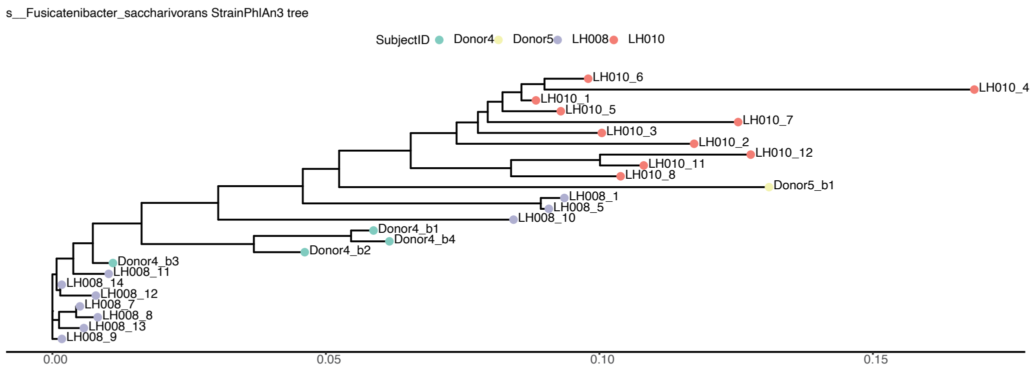
s__Fusicatenibacter_saccharivorans StrainPhlAn3 tree
SubjectID Donor4 Donor5 LH008 LH010
LH010_6
LH010_4
LH010_1
LH010_5
LH010_7
LH010_3
LH010_2
LH010_12
LH010_11
LH010_8
Donor5_b1
LH008_1
LH008_5
LH008_10
Donor4_b1
Donor4_b4
Donor4_b2
Donor4_b3
LH008_11
LH008_14
LH008_12
LH008_7
LH008_8
LH008_13
LH008_9
0.00
0.05
0.10
0.15

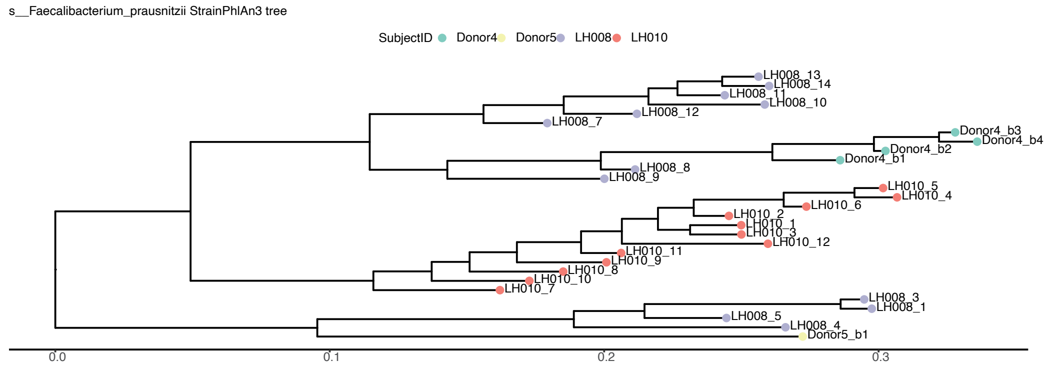
s__Faecalibacterium_prausnitzii StrainPhlAn3 tree
SubjectID Donor4 Donor5 LH008 LH010
LH008_13
LH008_14
LH008_11
LH008_10
LH008_12
LH008_7
Donor4_b3
Donor4_b4
Donor4_b2
Donor4_b1
LH008_8
LH008_9
LH010_5
LH010_4
LH010_6
LH010_2
LH010_1
LH010_3
LH010_12
LH010_11
LH010_9
LH010_8
LH010_10
LH010_7
LH008_3
LH008_1
LH008_5
LH008_4
Donor5_b1
0.0
0.1
0.2
0.3

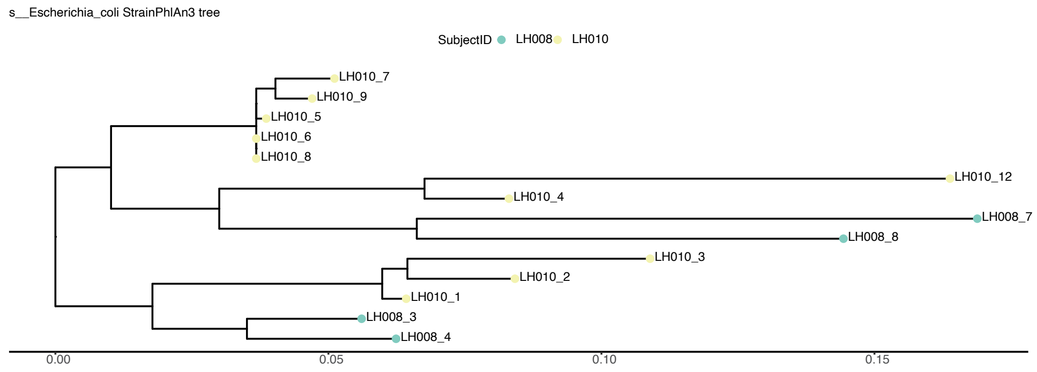
s__Escherichia_coli StrainPhlAn3 tree
SubjectID LH008 LH010
LH010_7
LH010_9
LH010_5
LH010_6
LH010_8
LH010_12
LH010_4
LH008_7
LH008_8
LH010_3
LH010_2
LH010_1
LH008_3
LH008_4
0.00
0.05
0.10
0.15

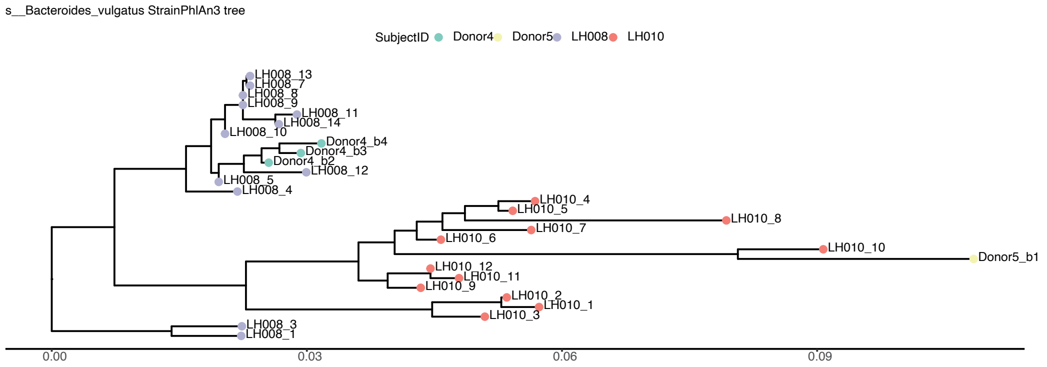
s__Bacteroides_vulgatus StrainPhlAn3 tree
SubjectID Donor4 Donor5 LH008 LH010
LH008_13
LH008_7
LH008_8
LH008_9
LH008_11
LH008_14
LH008_10
Donor4_b4
Donor4_b3
Donor4_b2
LH008_12
LH008_5
LH008_4
LH010_4
LH010_5
LH010_7
LH010_8
LH010_6
LH010_10
Donor5_b1
LH010_12
LH010_11
LH010_9
LH010_2
LH010_1
LH010_3
LH008_3
LH008_1
0.00
0.03
0.06
0.09

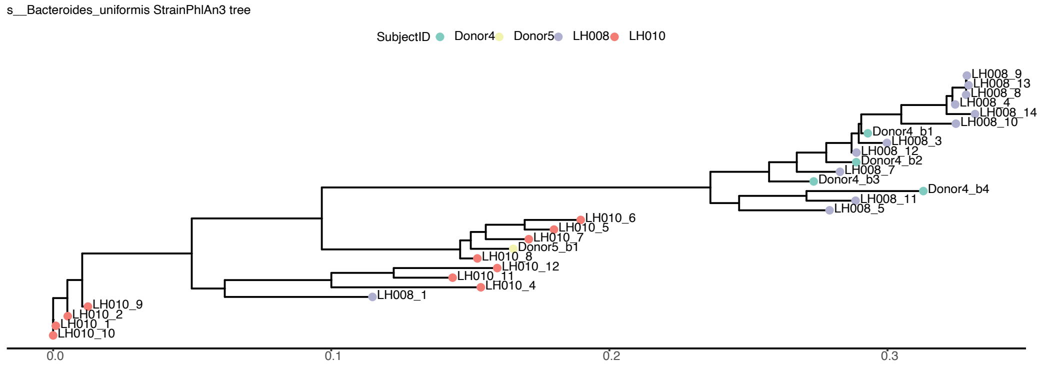
s__Bacteroides_uniformis StrainPhlAn3 tree
SubjectID Donor4 Donor5 LH008 LH010
LH008_9
LH008_13
LH008_8
LH008_4
LH008_14
LH008_10
Donor4_b1
LH008_3
LH008_12
Donor4_b2
LH008_7
Donor4_b3
Donor4_b4
LH008_11
LH008_5
LH010_6
LH010_5
LH010_7
Donor5_b1
LH010_8
LH010_12
LH010_11
LH010_4
LH008_1
LH010_9
LH010_2
LH010_1
LH010_10
0.0
0.1
0.2
0.3

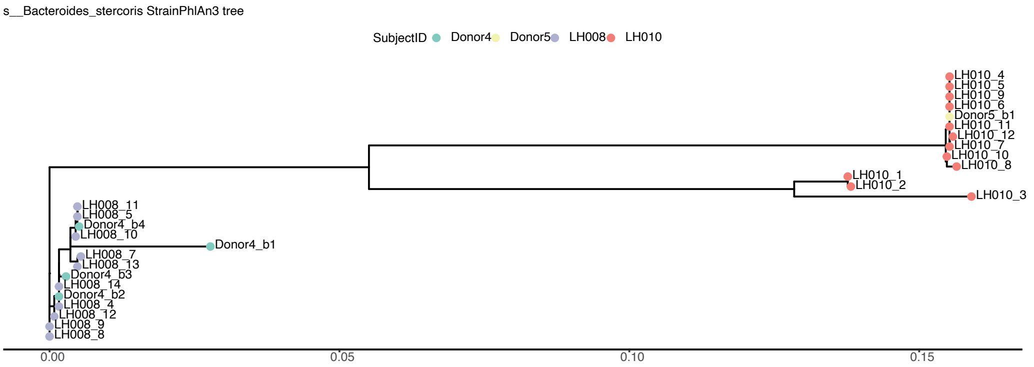
s__Bacteroides_stercoris StrainPhlAn3 tree
SubjectID Donor4 Donor5 LH008 LH010
LH010_4
LH010_5
LH010_9
LH010_6
Donor5_b1
LH010_11
LH010_12
LH010_7
LH010_10
LH010_8
LH010_1
LH010_2
LH010_3
LH008_11
LH008_5
Donor4_b4
LH008_10
Donor4_b1
LH008_7
LH008_13
Donor4_b3
LH008_14
Donor4_b2
LH008_4
LH008_12
LH008_9
LH008_8
0.00
0.05
0.10
0.15

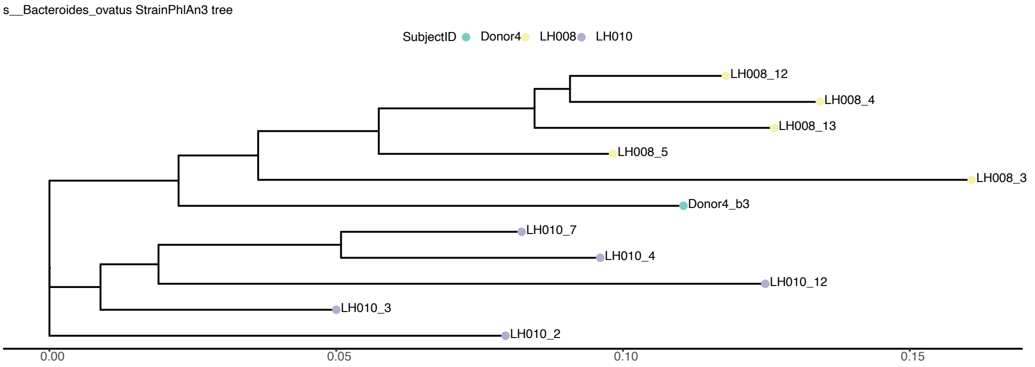
s__Bacteroides_ovatus StrainPhlAn3 tree
SubjectID Donor4 LH008 LH010
LH008_12
LH008_4
LH008_13
LH008_5
LH008_3
Donor4_b3
LH010_7
LH010_4
LH010_12
LH010_3
LH010_2
0.00
0.05
0.10
0.15

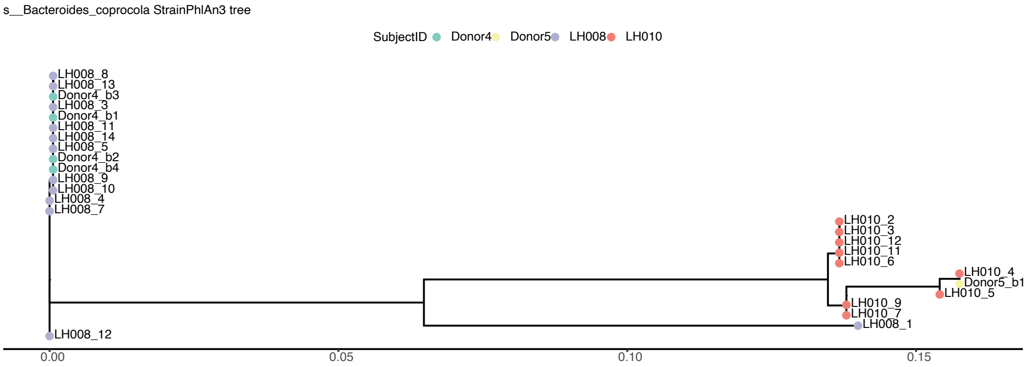
s__Bacteroides_coprocola StrainPhlAn3 tree
SubjectID Donor4 Donor5 LH008 LH010
LH008_8
LH008_13
Donor4_b3
LH008_3
Donor4_b1
LH008_11
LH008_14
LH008_5
Donor4_b2
Donor4_b4
LH008_9
LH008_10
LH008_4
LH008_7
LH010_2
LH010_3
LH010_12
LH010_11
LH010_6
LH010_4
Donor5_b1
LH010_5
LH010_9
LH010_7
LH008_1
LH008_12
0.00
0.05
0.10
0.15

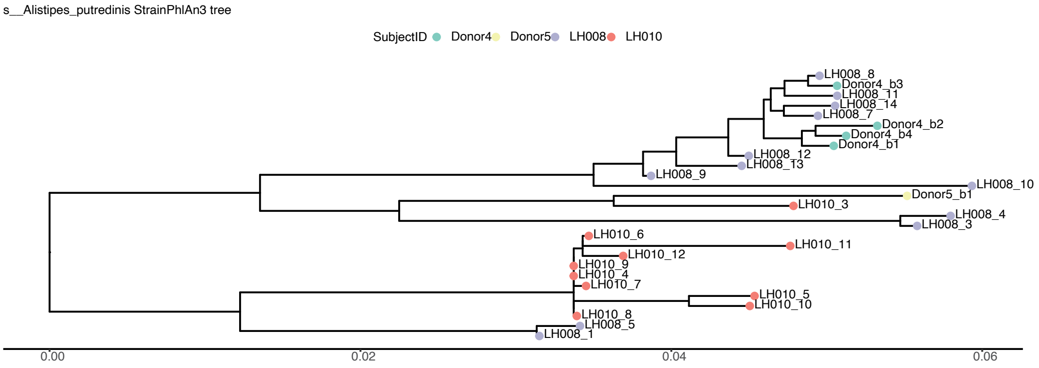
s__Alistipes_putredinis StrainPhlAn3 tree
SubjectID Donor4 Donor5 LH008 LH010
LH008_8
Donor4_b3
LH008_11
LH008_14
LH008_7
Donor4_b2
Donor4_b4
Donor4_b1
LH008_12
LH008_13
LH008_9
LH008_10
Donor5_b1
LH010_3
LH008_4
LH008_3
LH010_6
LH010_11
LH010_12
LH010_9
LH010_4
LH010_7
LH010_5
LH010_10
LH010_8
LH008_5
LH008_1
0.00
0.02
0.04
0.06

### Sup Fig5b

s__Eubacterium_rectale StrainPhlAn3 tree

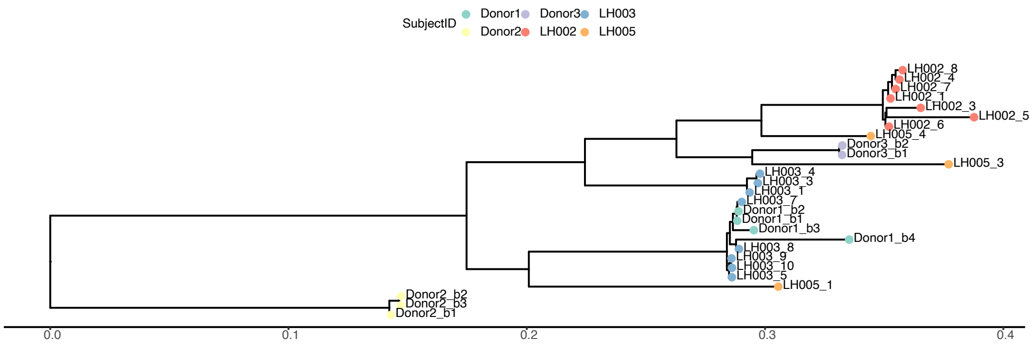

s__Cloacibacillus_porcorum StrainPhlAn3 tree

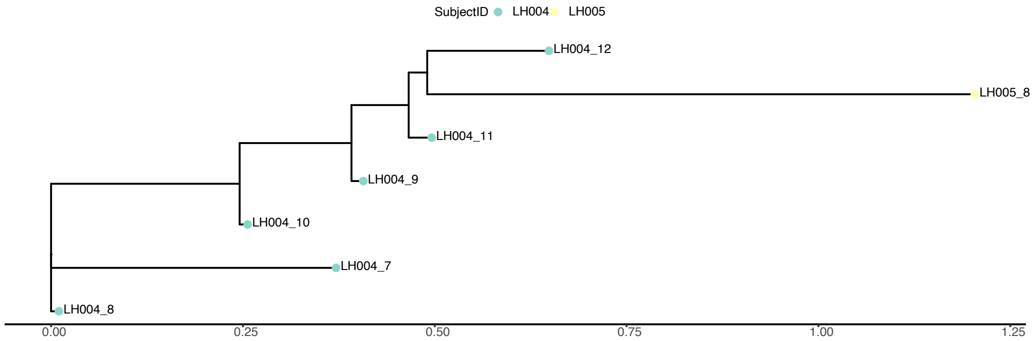

s__Bacteroides_dorei StrainPhlAn3 tree

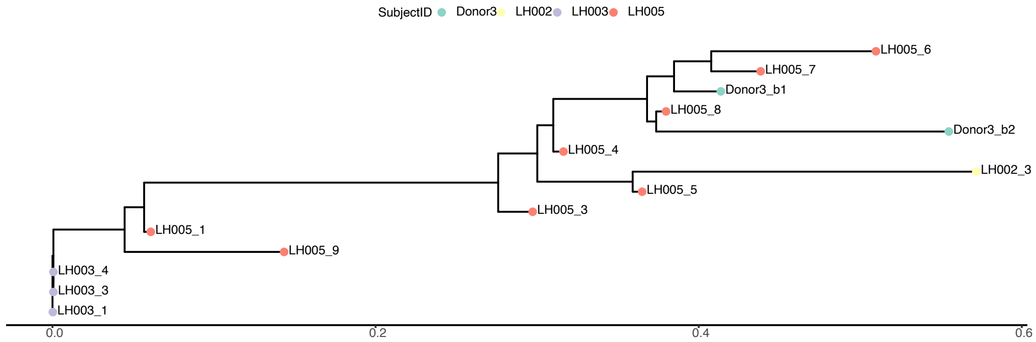

s__Bifidobacterium_dentium StrainPhlAn3 tree

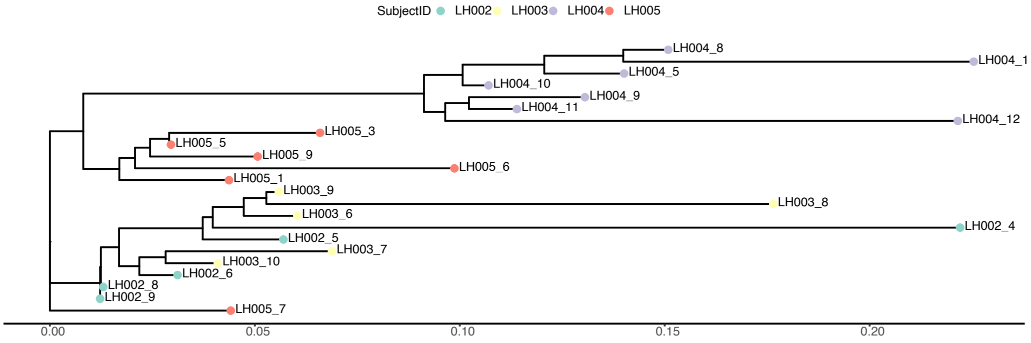

s__Oscillibacter_sp_57_20 StrainPhlAn3 tree

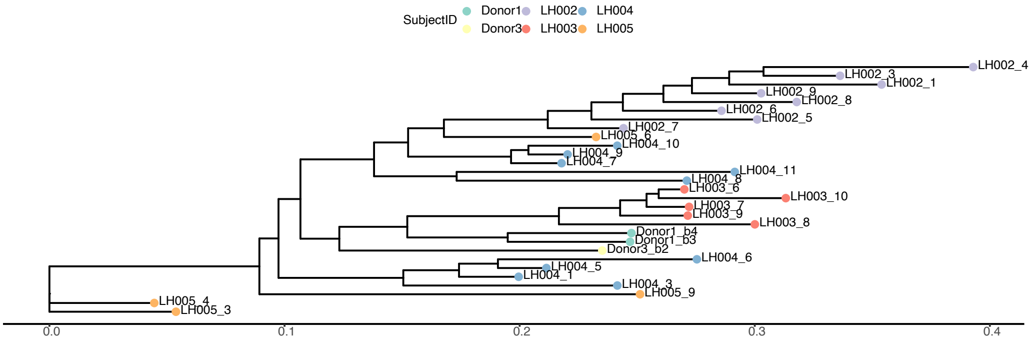

s__Intestinimonas_butyriciproducens StrainPhlAn3 tree

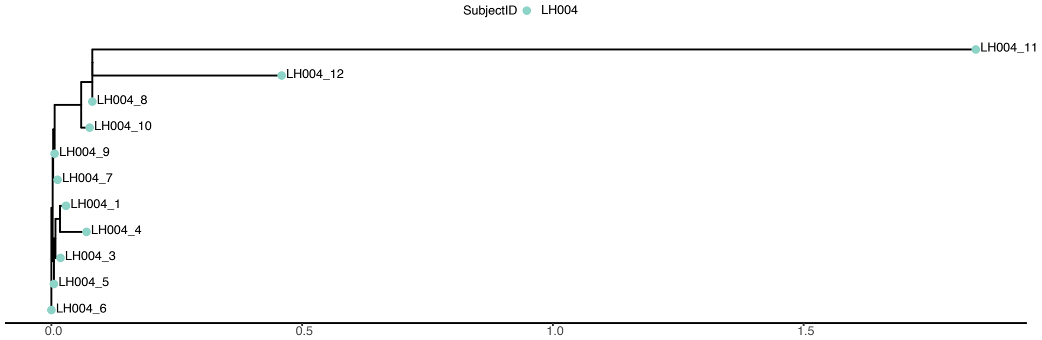

s__Bifidobacterium_adolescentis StrainPhlAn3 tree

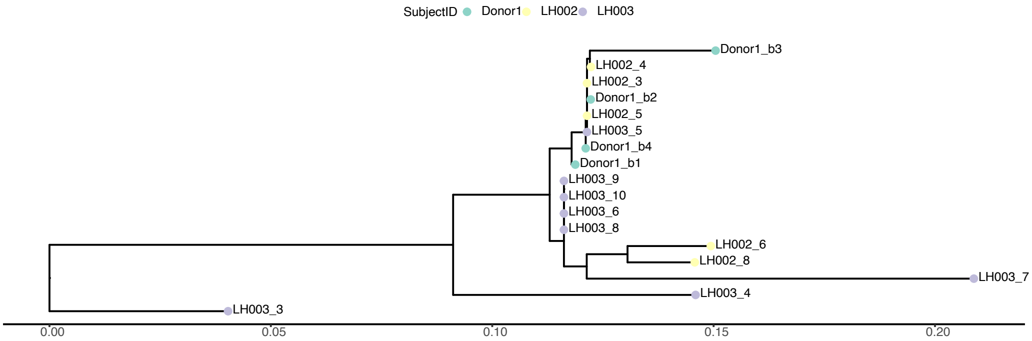

s__Parabacteroides_distasonis StrainPhlAn3 tree

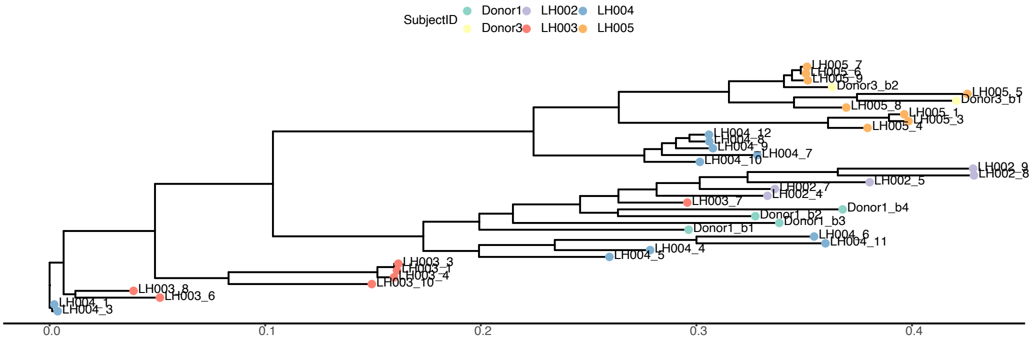

s__Ruminococcus_torques StrainPhlAn3 tree

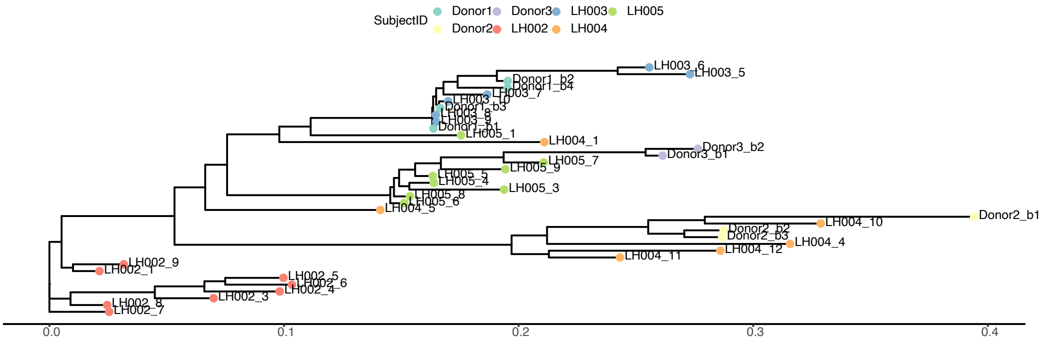

s__Roseburia_inulinivorans StrainPhlAn3 tree

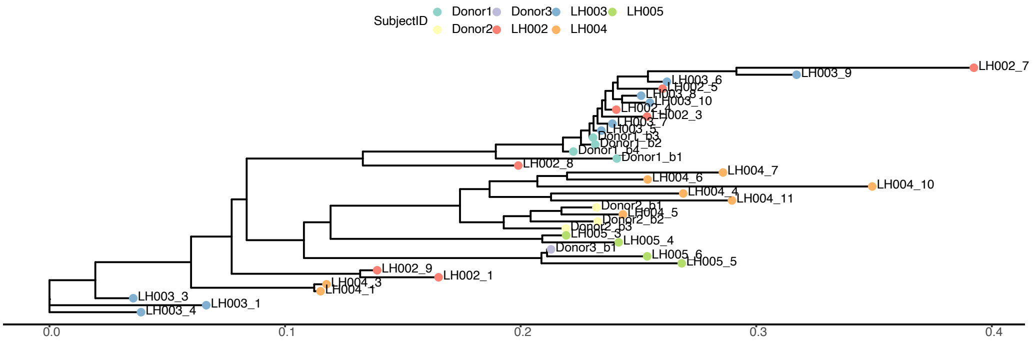

s__Klebsiella_pneumoniae StrainPhlAn3 tree

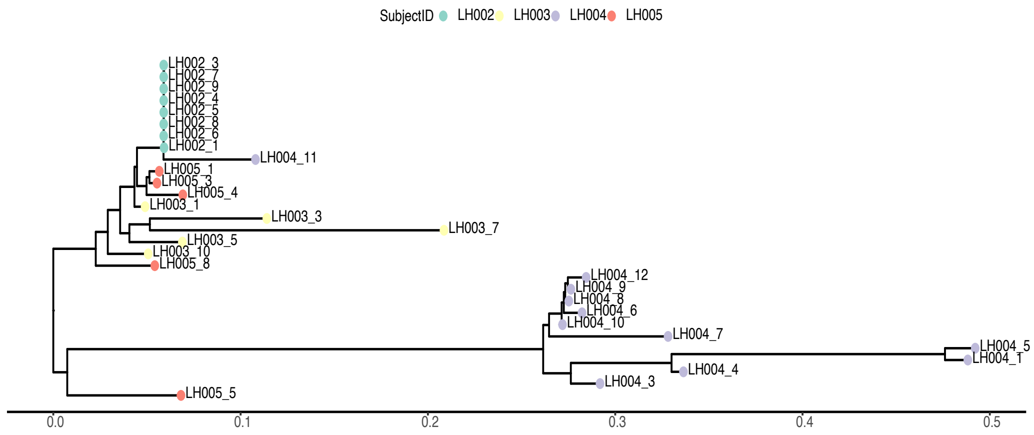

s__Eubacterium_hallii StrainPhlAn3 tree

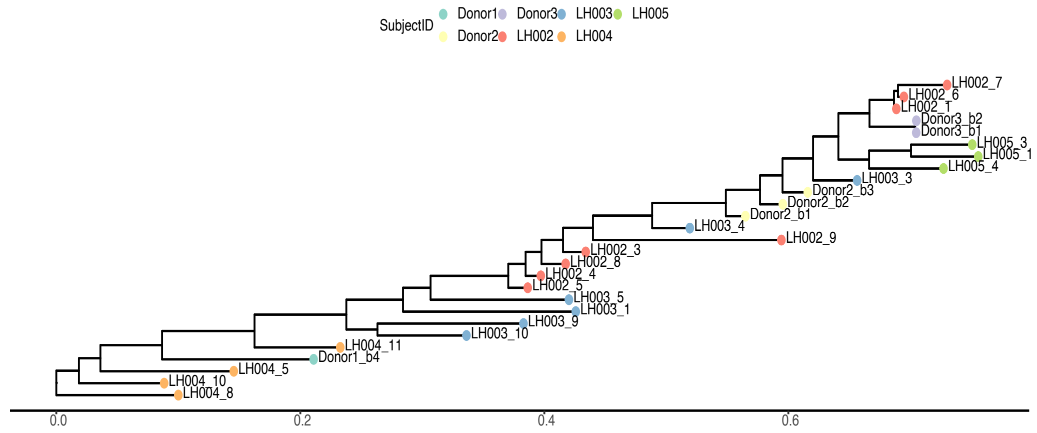

s__Akkermansia_muciniphila StrainPhlAn3 tree

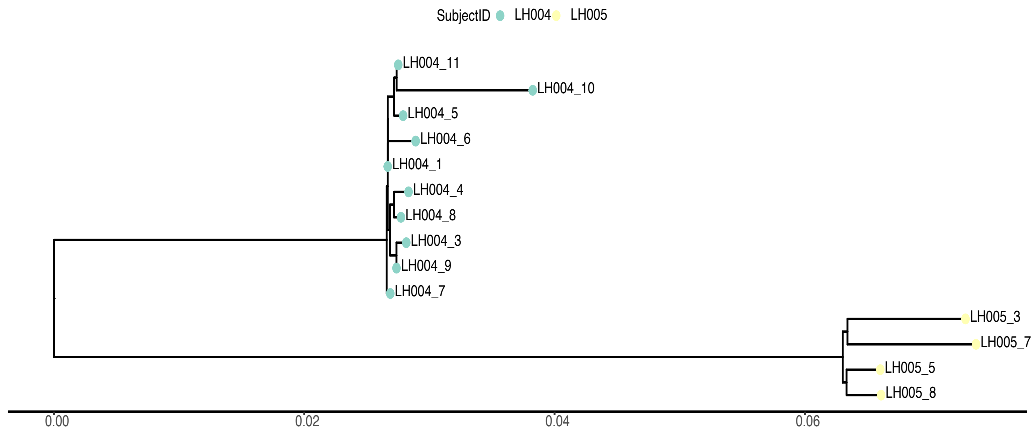

s__Parabacteroides_goldsteinii StrainPhlAn3 tree

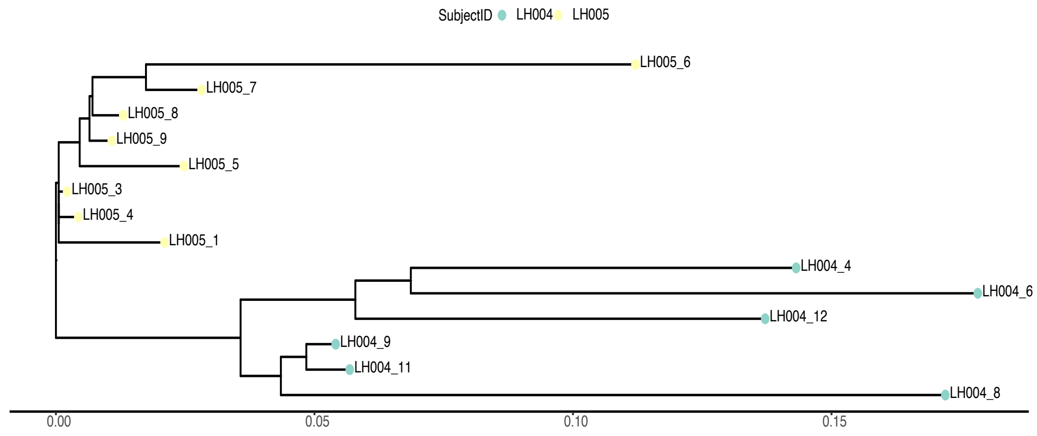

s__Bacteroides_fragilis StrainPhlAn3 tree

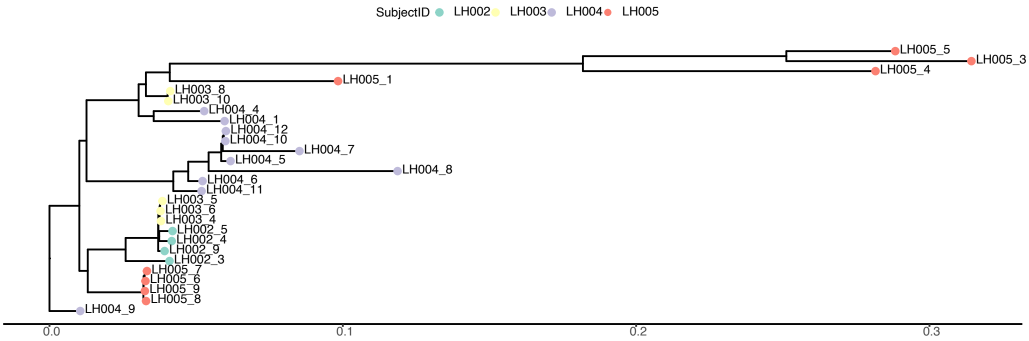

s__Collinsella_aerofaciens StrainPhlAn3 tree

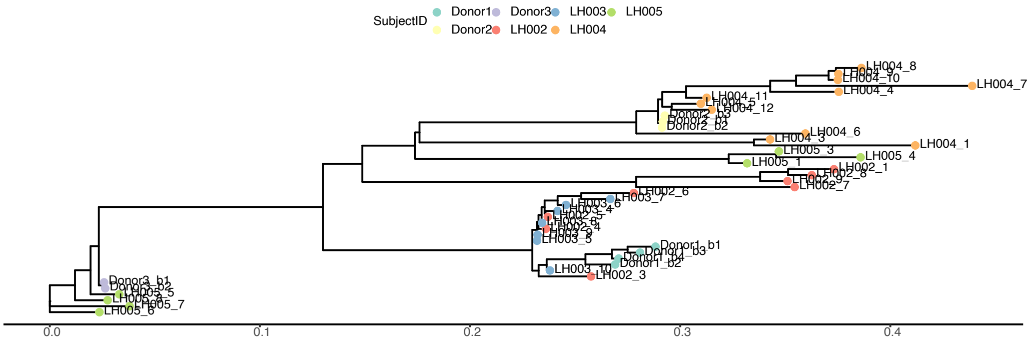

s__Prevotella_copri StrainPhlAn3 tree

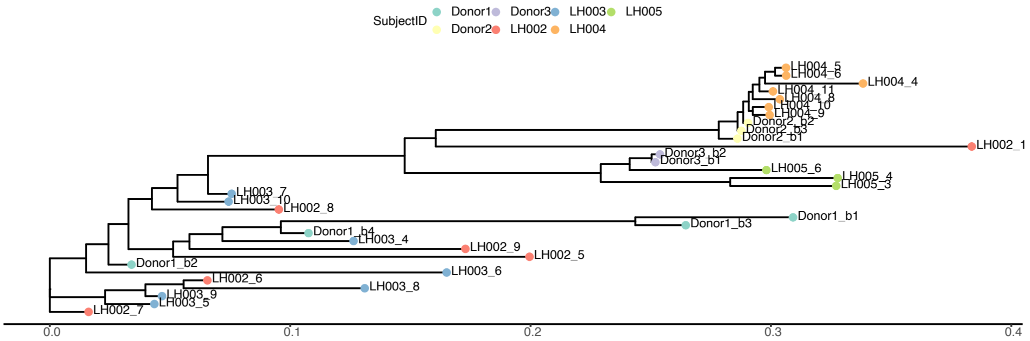

s__Klebsiella_quasipneumoniae StrainPhlAn3 tree

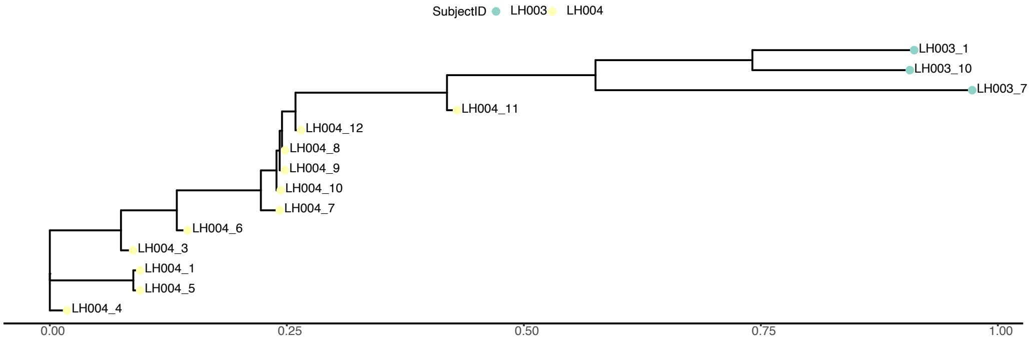

s__Phascolarctobacterium_succinatutens StrainPhlAn3 tree

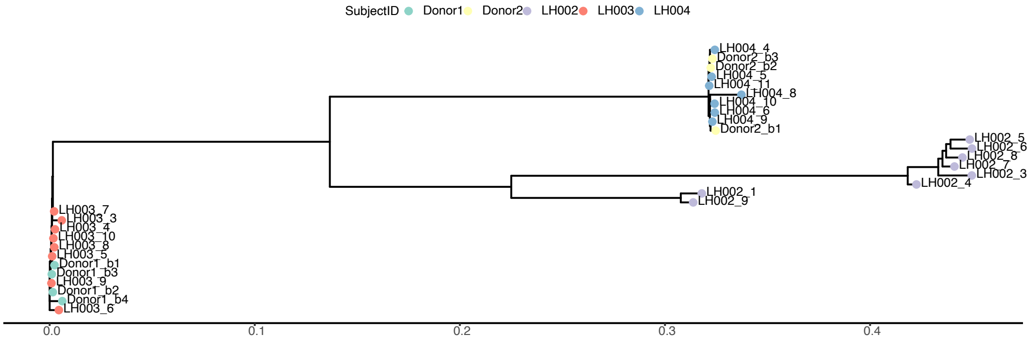

**Table S1**

**Table S1． Summary of All Adverse Events**

| Adverse Events | Subjects (%) | | | Occurrence | | |
|---|---|---|---|---|---|---|
| | Total | FMT monotherapy period | Combination therapy period | Total | FMT monotherapy period | Combination therapy period |
| At least one TEAE | 10 (100) | 4 (40·0) | 10 (100) | 87 | 9 | 78 |
| At least one grade 3 or above TEAE | 5 (50·0) | 2 (20·0) | 4 (40·0) | 8 | 2 | 6 |
| At least one early termination by TEAE | 0 | 0 | 0 | 0 | 0 | 0 |
| At least one treated TEAE | 6 (60·0) | 2 (20·0) | 6 (60·0) | 25 | 2 | 23 |
| At least one SAE | 2 (20·0) | 0 | 2 (20·0) | 2 | 0 | 2 |
| At least one TRAE | 3(30·0) | 1 (10·0) | 3 (30·0) | 4 | 2 | 4 |
| At least one grade 3 or above TRAE | 0 | 0 | 0 | 0 | 0 | 0 |
| At least one treated TRAE | 2 (20·0) | 0 | 2 (20·0) | 2 | 0 | 2 |
| At least one early termination by TRAE | 0 | 0 | 0 | 0 | 0 | 0 |
| At least one treatment related SAE | 0 | 0 | 0 | 0 | 0 | 0 |

**Table S2**

**Table S2． Severity of TEAE by SOC and PT (FMT monotherapy period)**

| SOC | All Grade | | Grade3 or above | | Grade 3 | |
|---|---|---|---|---|---|---|
| **PT** | **Occurrence** | **Subject (%)** | **Occurrence** | **Subject (%)** | **Occurrence** | **Subject (%)** |
| **At least one TEAE** | **9** | **4 (40·0)** | **2** | **2 (20·0)** | **2** | **2 (20·0)** |
| **Gastrointestinal disorders** | **4** | **3 (30·0)** | **1** | **1 (10·0)** | **1** | **1 (10·0)** |
| Vomiting | 2 | 2 (20·0) | 0 | 0 | 0 | 0 |
| Nausea | 1 | 1 (10·0) | 0 | 0 | 0 | 0 |
| Dysphagia | 1 | 1 (10·0) | 1 | 1 (10·0) | 1 | 1 (10·0) |
| **Investigations** | **3** | **2 (20·0)** | **0** | **0** | **0** | **0** |
| Conjugated bilirubin increased | 1 | 1 (10·0) | 0 | 0 | 0 | 0 |
| Blood urea increased | 1 | 1 (10·0) | 0 | 0 | 0 | 0 |
| Blood lactate dehydrogenase increased | 1 | 1 (10·0) | 0 | 0 | 0 | 0 |
| **Metabolism and nutrition disorders** | **1** | **1 (10·0)** | **0** | **0** | **0** | **0** |
| Hyponatremia | 1 | 1 (10·0) | 0 | 0 | 0 | 0 |
| **Musculoskeletal and connective tissue disorders** | **1** | **1 (10·0)** | **1** | **1 (10·0)** | **1** | **1 (10·0)** |
| Pain in extremity | 1 | 1 (10·0) | 1 | 1 (10·0) | 1 | 1 (10·0) |

**Table S3**

**Table S3． Severity of TEAE by SOC and PT (anti-PD-1 and FMT combination therapy period)**

| SOC | All Grade | | Grade 3 or above | | Grade 3 | |
|---|---|---|---|---|---|---|
| PT | Occurrence | Subject (%) | Occurrence | Subject (%) | Occurrence | Subject (%) |
| **At least one TEAE** | **78** | **10 (100·0)** | **6** | **4 (40·0)** | **5** | **4 (40·0)** |
| **Gastrointestinal disorders** | **14** | **6 (60·0)** | **2** | **2 (20·0)** | **2** | **2 (20·0)** |
| Nausea | 3 | 3 (30·0) | 0 | 0 | 0 | 0 |
| Abdominal pain | 5 | 2 (20·0) | 1 | 1 (10·0) | 1 | 1 (10·0) |
| Dysphagia | 2 | 2 (20·0) | 1 | 1 (10·0) | 1 | 1 (10·0) |
| Constipation | 1 | 1 (10·0) | 0 | 0 | 0 | 0 |
| Diarrhea | 1 | 1 (10·0) | 0 | 0 | 0 | 0 |
| Abdominal distention | 1 | 1 (10·0) | 0 | 0 | 0 | 0 |
| Vomiting | 1 | 1 (10·0) | 0 | 0 | 0 | 0 |
| **Metabolism and nutrition disorders** | **6** | **2 (20·0)** | **0** | **0** | **0** | **0** |
| Hyponatremia | 2 | 2 (20·0) | 0 | 0 | 0 | 0 |
| Hyperuricemia | 2 | 2 (20·0) | 0 | 0 | 0 | 0 |
| Hypoglycemia | 1 | 1 (10·0) | 0 | 0 | 0 | 0 |
| Decreased appetite | 1 | 1 (10·0) | 0 | 0 | 0 | 0 |
| **Investigations** | **31** | **9 (90·0)** | **0** | **0** | **0** | **0** |
| Weight loss | 9 | 6 (60·0) | 0 | 0 | 0 | 0 |
| White blood cell decreased | 4 | 2 (20·0) | 0 | 0 | 0 | 0 |
| Conjugated bilirubin increased | 2 | 2 (20·0) | 0 | 0 | 0 | 0 |
| Urine white blood cell positive | 5 | 2 (20·0) | 0 | 0 | 0 | 0 |
| Blood lactate dehydrogenase increased | 2 | 2 (20·0) | 0 | 0 | 0 | 0 |
| Triiodothyronine decreased | 2 | 1 (10·0) | 0 | 0 | 0 | 0 |
| Blood thyroid stimulating hormone decreased | 1 | 1 (10·0) | 0 | 0 | 0 | 0 |
| Blood thyroid stimulating hormone increased | 1 | 1 (10·0) | 0 | 0 | 0 | 0 |
| Blood bilirubin increased | 1 | 1 (10·0) | 0 | 0 | 0 | 0 |
| Blood creatine phosphokinase increased | 1 | 1 (10·0) | 0 | 0 | 0 | 0 |
| Blood urea increased | 1 | 1 (10·0) | 0 | 0 | 0 | 0 |
| Platelet count decreased | 1 | 1 (10·0) | 0 | 0 | 0 | 0 |
| Free triiodothyronine decreased | 1 | 1 (10·0) | 0 | 0 | 0 | 0 |
| **Blood and Lymphatic System Disorders** | **9** | **6 (60·0)** | **0** | **0** | **0** | **0** |
| Anemia | 9 | 6 (60·0) | 0 | 0 | 0 | 0 |
| **Infections and infestations** | **2** | **2 (20·0)** | **1** | **1 (10·0)** | **1** | **1 (10·0)** |
| COVID-19 | 1 | 1 (10·0) | 0 | 0 | 0 | 0 |
| Infectious pneumonia | 1 | 1 (10·0) | 1 | 1 (10·0) | 1 | 1 (10·0) |
| **Musculoskeletal and connective tissue disorders** | **5** | **2 (20·0)** | **1** | **1 (10·0)** | **1** | **1 (10·0)** |
| Back pain | 5 | 2 (20·0) | 1 | 1 (10·0) | 1 | 1 (10·0) |
| **Psychiatric disorders** | **2** | **2 (20·0)** | **0** | **0** | **0** | **0** |
| Insomnia | 2 | 2 (20·0) | 0 | 0 | 0 | 0 |
| **General disorders and administration site conditions** | **4** | **2 (20·0)** | **2** | **1 (10·0)** | **1** | **1 (10·0)** |
| Fever | 2 | 1 (10·0) | 0 | 0 | 0 | 0 |
| Fatigue | 1 | 1 (10·0) | 1 | 1 (10·0) | 1 | 1 (10·0) |
| Disease progression | 1 | 1 (10·0) | 1 | 1 (10·0) | 0 | 0 |
| **Renal and urinary disorders** | **4** | **2 (20·0)** | **0** | **0** | **0** | **0** |
| Proteinuria | 4 | 2 (20·0) | 0 | 0 | 0 | 0 |
| **Respiratory, thoracic and mediastinal disorders** | **1** | **1 (10·0)** | **0** | **0** | **0** | **0** |
| Allergic rhinitis | 1 | 1 (10·0) | 0 | 0 | 0 | 0 |

**Table S4**

**Table S4． Severity of TRAE by SOC and PT (FMT monotherapy period)**

| SOC | All Grade | | Grade 3 or above | | Grade 3 | |
|---|---|---|---|---|---|---|
| PT | Occurrence | Subject (%) | Occurrence | Subject (%) | Occurrence | Subject (%) |
| **At least one TRAE** | **2** | **1 (10·0)** | **0** | **0** | **0** | **0** |
| **Gastrointestinal disorders** | **2** | **1 (10·0)** | **0** | **0** | **0** | **0** |
| Vomiting | 1 | 1 (10·0) | 0 | 0 | 0 | 0 |
| Nausea | 1 | 1 (10·0) | 0 | 0 | 0 | 0 |

**Table S5**

**Table S5． Severity of TRAE by SOC and PT ((anti-PD-1 and FMT combination therapy period)**

| SOC | All Grade | | Grade3 or above | | Grade 3 | |
|---|---|---|---|---|---|---|
| PT | Occurrence | Subject (%) | Occurrence | Subject (%) | Occurrence | Subject (%) |
| **At least one TRAE** | 4 | 3 (30·0) | 0 | 0 | 0 | 0 |
| **Gastrointestinal disorders** | 4 | 3 (30·0) | 0 | 0 | 0 | 0 |
| Nausea | 3 | 3 (30·0) | 0 | 0 | 0 | 0 |
| Constipation | 1 | 1 (10·0) | 0 | 0 | 0 | 0 |

**Table S6**

**Table S6 . Comparison of Metacyc pathways between Rs and NRs after FMT using linear mixed effect model**

| PathwayIDs | NR(n=35) mean(sd) | R(n=13) mean(sd) | pvalue | adjusted·pvalue |
|---|---|---|---|---|
| PWY-7312 | 3·56E-05±5·99E-05 | 6·40E-04±5·75E-04 | 6·04E-08 | 4·64E-05 |
| PWY-6507 | 1·43E-03±4·65E-04 | 2·14E-03±4·85E-04 | 2·77E-05 | 0·01 |

**Table S7**

**Table S7． List of patients’ and donors’ samples in the downstream analysis**

| SujectID | cancer_group | baseline_sample_number | post_sample_number | total_sample_number | clinical_outcome | fmt_donor |
|---|---|---|---|---|---|---|
| LH001 | CRC | 1 | 6 | 7 | NR | Donor1 |
| LH002 | GC | 1 | 7 | 8 | NR | Donor1 |
| LH003 | GC | 1 | 8 | 9 | NR | Donor1 |
| LH004 | GC | 1 | 10 | 11 | NR | Donor2 |
| LH005 | GC | 1 | 7 | 8 | NR | Donor3 |
| LH006 | GC | 1 | 11 | 12 | PRR | Donor2 |
| LH007 | GC | 1 | 12 | 13 | PRR | Donor4 |
| LH008 | GC | 1 | 11 | 12 | R | Donor4 |
| LH009 | CRC | 1 | 10 | 11 | NR | Donor4 |
| LH010 | GC | 1 | 11 | 12 | R | Donor5 |
| Donor1 | | | | 4 | ineffective | |
| Donor2 | | | | 3 | ineffective | |
| Donor3 | | | | 2 | ineffective | |
| Donor4 | | | | 4 | effective | |
| Donor5 | | | | 1 | effective | |

**Table S8**

**Table S8． Mean ± SEM values of clinical outcome by subject response at baseline**

| project name | NR(n=4) | PRR(n=2) | R(n=2) |
|---|---|---|---|
| CD4+ % CD3+ cells | 55·00±17·60 | 62·00±4·31 | 71·4±7·07 |
| CCR7+CD45RA- % CD4+ cells | 12·30±8·11 | 29·20±3·04 | 35·6±2·83 |
| Foxp3+ % CD4+ cells | 0·21±0·10 | 0·11±0·06 | 0·03±0·02 |
| Ki67+ % CD3+ cells | 0·21±0·30 | 1·14±0·06 | 0·6±0·15 |
| CD279+ % lymphocytes | 10·50±3·91 | 12·00±2·83 | 11·3±5·23 |
| TCR | 10·40±1·54 | 12·6±0·65 | 14·61±0·38 |

**Table S9**

**Table S9． Mean ± SEM values of clinical outcome by subject response at post**

| project name | NR(n=9) | PRR(n=6) | R(n=6) |
|---|---|---|---|
| CD4+ % CD3+ cells | 53·50±18·00 | 61·90±12·30 | 71·10±9·89 |
| CCR7+CD45RA- % CD4+ cells | 22·70±9·46 | 33·50±6·10 | 38·3±4·96 |
| Foxp3+ % CD4+ cells | 0·23±0·44 | 0·04±0·04 | 0·05±0·04 |
| Ki67+ % CD3+ cells | 0·05±0·05 | 0·58±0·29 | 0·41±0·50 |
| CD279+ % lymphocytes | 9·24±11·10 | 13·00±18·7 | 5·73±8·57 |
| TCR | 10·30±1·50 (n=5) | 12·8±0·21 (n=3) | 14·3±0·60 (n=4) |

#### METHODS SUPPLEMENT

##### Study participants

This study (NCT04130763) is an open label, single-arm, single center, Phase 1 study that included patients with advanced GI cancer who were resistant or refractory to the ICI treatment. Resistant or refractory to ICI treatment was defined as patients that must have received at least 2 doses of ICI treatment previously but reached disease progression on the treatment, determined by iRECIST criteria prior to enrollment into the study. Previous treatments with surgery, radiation therapy, chemotherapy, or targeted therapy, such as HER2 inhibitors, were allowed. Patients recruited were at least 18 years of age, with histologically confirmed GI cancer. At least 3 months of estimated survival duration and an ECOG score of 0-1 was required. Major exclusion criteria included: a history of autoimmune diseases; active GI diseases; unstable brain metastases; the use of daily corticosteroids >10 mg (prednisone or equivalent); a condition that must use antibiotics during the study.

##### Study enrollment

Ten patients were enrolled and treated at a single center in Beijing, China, out of 11 GI cancer patients who were screened against inclusion and exclusion criteria. The research was carried out under the approval of Medical Ethics Committee of Beijing Cancer Hospital and in accordance with the applicable regulatory requirements of National Health Commission of the People's Republic of China; the International Council for Harmonization, Guideline for Good Clinical Practice (ICH GCP); and the principles of the Declaration of Helsinki. All patients provided written informed consent and were able to withdraw consent at any time without compromising their treatment for any of their medical conditions.

##### Inclusion Criteria

1. Patient had a histologically or cytologically confirmed diagnosis of unresectable or metastatic solid tumors originating from the GI tract.
2. Patient was able and willing to provide pathological tissue embedded in wax blocks or paraffin sections.
3. Patient had received any number of radiation therapy, chemotherapy, vaccine therapy or other oncological therapy are permitted.
4. Patient was receiving or has received at least 2-dose injections of systemic PD-(L)1 immunotherapy, and imaging results confirmed progressive disease (PD). According to iRECIST, PD is defined as an increase in the length of the lesion > 20% or occurrence of new lesion or non-target lesion progression. Anti-PD-(L)1 drugs can contain pembrolizumab, nivolumab or any other anti-PD-(L)1 drugs that have passed phase 2 clinical development. Patients are eligible when the previous failed anti-PD-(L)1 treatment was initiated within one year of the first dose of this trial.

5. Patient was willing and able to swallow at least 20 FMT capsules.

6. Patient was willing and able to sign the informed consent form.

7. Patient consented to receive follow-up medical imaging to determine disease progression and provide stool samples before and after taking capsules at each follow-up visit.

8. Patient was at least 18 years old, male or female.

9. Patient had an ECOG performance of 0 or 1.

10. For women with childbearing potential, the results of blood pregnancy test within 7 days prior to enrollment or the results of urine pregnancy test within 72 hours prior to enrollment had to be negative.

11. Patient must have had basic body function. Blood test results needed to reach the following indexes: Absolute neutrophil count (ANC)≥1500/mcL, platelets≥100,000/mcL, hemoglobin≥9 g/dL or 5.6 mmol/L, no transfusion or EPO dependency (within 7 days of assessment), serum creatinine≤1.5×upper limit of normal (ULN) or creatinine clearance≥60 mL/min, serum total bilirubin≤1.5×ULN or direct bilirubin≤ULN, AST (SGOT) and ALT (SGPT)≤2.5×ULN for patients with serum total bilirubin >1.5 ULN or ≤5×ULN for patients with liver metastases, albumin≥2.5 mg/dL, coagulation indexes INR or PT ≤1.5×ULN. Unless patient was receiving anticoagulant therapy, coagulation indexes were within normal range of therapy.

12. Expected survival duration ≥3 months.

**Exclusion Criteria**

1. Patient with irritable bowel syndrome, toxic megacolon, and severe dietary allergies (including severe allergic to shellfish, nuts, seafood).

2. Patient had responded to anti-PD-(L)1 therapy or had a stable disease status (per iRECIST at CR, PR, or SD).

3. Patient had participated in any other clinical trial within 4 weeks before the first dose of FMT capsule treatment.

4. Patient had highly severe symptoms including rapidly declining ECOG performance; rapidly worsening symptoms; lesion transferring to critical sites and requiring urgent medical intervention.

5. Patient had a known history of malignant blood diseases, primary brain tumor or sarcoma, or other primary solid tumors except gastrointestinal tumors.

6. Patient had progressing CNS metastases or leptomeningeal metastases. Patients with stable brain metastases had to be re-screened by brain MRI or CT scan within two weeks before enrollment to ensure no disease

progression, and simultaneously take ≤10mg steroid daily one week prior to treatment. Patients with no history of CNS metastases and no signs of CNS metastases did not need another medical imaging examination for brain diseases.

7. Patient had a severe hypersensitivity or reaction to anti-PD-(L)1 immunotherapy.
8. Patient had an autoimmune disease or a history of autoimmune disease or required treatment with systemic steroid (prednisone >10 mg daily or equivalent dose of similar drugs) or immunosuppressive agents. The following cases were exceptions: local, ophthalmic, intra-articular, intranasal, or inhaled corticosteroids with extremely low systemic absorption; hormone replacement therapy; short-term (≤7 days) treatment of corticosteroids for preventative use (e.g., allergy to contrast agents).
9. Patient had pneumonitis or had a history of (non-infectious) pneumonitis requiring steroid therapy.
10. Patient had severe cardiovascular disease (e.g., drug-uncontrolled congestive heart failure, hypertension, cardiac ischemia, myocardial infarction, and severe cardiac arrhythmia), bleeding disorders, severe obstructive or restrictive pulmonary diseases, or systemic infections.
11. Patient had active human immunodeficiency virus (HIV) infection (performance as HIV 1/2 antibodies and/or positive).
12. Patient had active Hepatitis B (HBV) or Hepatitis C (HCV) infection.
13. Patient had known active tuberculosis.
14. Patient had received a live vaccine or live attenuated vaccine within 4 weeks prior to enrollment.
15. Patient had reported adverse events (per CTCAE 5.0, ≥grade 2) due to drug treatment within 4 weeks and not recovered from it. Patients who had undergone major surgery had to have completely recovered from toxicity and complications of previous interventions prior to participating in the study.
16. Patient had received anti-tumor therapy except experimental drugs (e.g., chemotherapy, targeted small molecule therapy or radiotherapy) within 2 weeks prior to screening or planned to receive anti-tumor therapy except experimental drugs (e.g., chemotherapy, targeted small molecule therapy or radiotherapy) during the study period. Radiotherapy used for pain control was an exception.
17. Patient had a known history of psychiatric disorders or drug abuse.

18. Patient was pregnant or breastfeeding, or subjects (including male subject and his female spouse) could not take effective contraceptive measures at the time of signing the informed consent until 120 days after the final anti-PD-1 therapy combined with FMT treatment.
19. Patient could not stop antibiotics treatment 24 hours before administration due to infection, etc.
20. Other cases that investigators qualified as relevant for patients who could not participate in the study, e.g., any medical history, treatment history, or history of abnormal test data that possibly confuses the study results, or interferes with patients' participation in the whole study, or damages patient interests.

**Fecal microbiota transplantation (FMT)**

With careful consideration of the safety and wellbeing of the study participants who were GI cancer patients with primary cancer lesions at GI track and at least half of them had GI surgery, no gut preparations were performed. FMT monotherapy was orally performed via capsules that each contained 0.651-0.690g (average amount by weight =0.676g) of fecal extract from strictly screened healthy donors. Each patient received 60 capsules administered in three doses over three to five days during the first week of the study treatment. The anti-PD-1+FMT therapy started at the second week. An FMT maintenance treatment (mFMT, a dose at 10 capsules per treatment) combined with Nivolumab (3mg/kg, q14d) at every cycle of the anti-PD-1 treatment until the patient completed study or early terminated from the study. If a patient benefitted from the anti-PD-1+FMT therapy, at least 3 doses of mFMT were provided to patients during the extension phase for the purpose of strengthening the colonization of gut bacteria.

**Combination treatment**

The anti-PD-1+FMT therapy was administered every other week after FMT monotherapy. Total on-study treatment duration was 12 weeks for 6 cycles of the anti-PD-1+FMT therapy. At the end of the 6 cycles of therapy, patient responding to the anti-PD-1+FMT therapy continued 2-3 more cycles of the treatment in an extended study phase, followed by anti-PD-1 monotherapy, with mFMT as needed according to the investigator. Patients were allowed to switch to another anti-PD-1 agent if they could not afford nivolumab. One of the patients switched to Sintilimab (200mg, q21d) started from extension phase.

**Sample and data collection**

Stool samples were collected at each study visit (baseline, weekly for Week 1-3, every other week for Week 4-12, end of the study, and study extension phase) for gut microbiome analysis. Peripheral blood was collected for T cell Receptor analysis and flow cytometric analysis at the baselines (before FMT monotherapy and anti-PD-1+FMT therapy), mid-term evaluation, and end of the study visit. Tumor tissues (in the form of pathologic slices) were also collected at the screening visit for the analysis of Microsatellite Instability (MSI), Tumor Mutational Burden (TMB), and PD-L1 expression. Safety related blood and urine samples were collected at baseline, prior to anti-PD-1+FMT dosing and at the end of the study visit.

**Safety evaluation**

FMT and anti-PD-1-related toxicities were evaluated using Common Terminology Criteria for Adverse Events (CTCAE) 5.0 grading scale. FMT-specific toxicities were collected during FMT monotherapy period. Adverse events occurred after the start of the combination therapy were evaluated separately and jointly, i.e., whether the event was related only to FMT, only to Nivolumab, or possibly to both. The safety evaluation was carried out continuously throughout the study in both treatment phases, FMT monotherapy phase and anti-PD-1+FMT therapy phase. Any patient who experienced a serious adverse event had the anti-PD-1+FMT therapy suspended until resolved, stable, or discontinued at the discretion of the treating oncologist. All adverse events were reviewed and monitored to ensure consistency and accuracy.

**Efficacy valuation**

iRECIST was used as main efficacy evaluation criteria for all imaging evaluations. Per protocol, a responder (R) is defined as a patient who is not icPD or considered clinical progression per efficacy evaluation at the first and the end of the study evaluation after 12 weeks of the anti-PD-1+FMT therapy. A partial responder (PRR) is defined as a patient who has SD or better efficacy outcome at the first efficacy evaluation (after 6 weeks of treatment) but has icPD or clinical progression after another 4-8 weeks of anti-PD-1+FMT treatments. A patient will also be considered a PRR if the outcome at the first efficacy evaluation is iuPD without clinical progression, followed by an iSD at the confirmation evaluation (additional 4-8 weeks of treatment), but eventually reached icPD or clinical progression without further improvement. Non-responder (NR) is defined as a patient who had icPD or iuPD with signs of clinical progression at either the first or the end of study evaluation. Efficacy evaluation was performed in a frequency of every 2-4 cycles of the anti-PD-1+FMT therapy.

**Outcomes**

The primary outcome was ORR by iRECIST criteria and the safety of FMT administration in advanced GI cancer patients who were resistant to anti-PD-1 therapy. Secondary outcome included safety of anti-PD-1+FMT therapy in treating advanced GI cancer patients, the changes in patient gut microbiome composition and the changes in peripheral immune profiles.

**Healthy stool donor inclusion and exclusion criteria**

Donor Qualification: FMT capsules (XBI-302) were provided by Shenzhen Xbiome Biotech Co.,LTD. All FMT capsules used in this study were produced in between 2019–2022. FMT capsule production was conducted under compliance with GMP. Donors were selected from healthy volunteers who must pass the donor qualification process: completion of the screening questionnaires truthfully; completion of an in-person interview; completion of all required laboratory tests; and tested negative on all GI pathogen detection tests.

**Key inclusion criteria:**

1. Aged between 18-40 years old.
2. Had normal BMI (18-23.9 $Kg/m^2$) according to Chinese standard.
3. Had regular sleeping hours with enough sleep, healthy eating habits, and regular exercise.
4. Monogamous sexual relationship with same sex partner.
5. Did not work in Senior centers, hospitals.
6. Never had organ transplantation, blood infusion (within previous 6 months) or dialysis.

**Key exclusion criteria:**

1. A history of diagnosed gastrointestinal disorders, liver disorders or cancers, including but not limited to: gastroesophageal reflux; peptic ulcer disease; celiac disease; or inflammatory bowel disease (Crohn's disease or ulcerative colitis).
2. History of severe allergy or asthma.
3. Family history or self-medical history of auto-immune diseases, diabetes, hypertension, metabolic syndrome, or familial malignancy.
4. History of psychiatric disorders (major affective disorder, psychotic illness or ongoing use of any psychiatric medications).
5. Symptoms of diarrhea, constipation, or blood in stool within 3 months.
6. Any underlying metabolic disease including hypertension, hyperlipidemia, diabetes, metabolic syndrome cirrhosis, or nonalcoholic fatty liver disease.
7. Any of the following symptoms 14 days before stool collection: fever, cough, loss of taste, anosmia, fatigue/malaise, myalgia, sore throat, nausea, abdominal pain, diarrhea, dyspnea, chest pain, rash conjunctivitis, or headache.
8. Any symptoms suggestive of SARS-CoV-2 or known SARS-CoV-2 infection within the previous 3 months.
9. Previous surgery on the intestine, liver or gallbladder.

10. Recent travel to a pandemic region of any pathogen, tropic area, and the regions with high risk for infectious diseases.

11. Use of any antibiotics, steroids, immunosuppressant, or weight loss medication within the previous 6 months. Continuously use proton pump inhibitors for more than 30 days within the previous 6 months.

12. Hospitalization for more than 2 days within the previous 3 months or more than 5 days within the previous 12 months.

13. History of drug abuse.

14. Fecal occult blood (FOB) positive; Abnormal fecal Calprotectin.

15. Any positive laboratory results for a transmissible pathogen.

16. Positive detection of the following pathogens on screening laboratory testing: *Shigella, Salmonella sp., Yersinia, Campylobacter, sorbitol-negative E. coli 0157-H7, Plesiomonas, enteropathogenic E. coli* (EPEC), Shiga toxin-producing *E. coli* (STEC), *Shigella/Enteroinvasive E. coli* (EIEC), *candida albicans, vibrio, C. difficile,* or *Helicobacter pylori.*

17. Positive stool test for any multidrug-resistant organisms (extended spectrum β--lactamase (ESBL)-producing *enterobacteriaceae*, vancomycin-resistant *enterococci* (VRE), carbapenem-resistant *enterobacteriaceae* (CRE/CPE), or methicillin-resistant *staphylococcus aureus* (MRSA)).

18. Positive detection for norovirus, rotavirus, adenovirus, sapovirus, astrovirus, enteroviruses A71, coxsackie A16, angiostrongylus cantonensis and strongyloides stercoralis, parasites and egg cells.

19. Positive detection for human immunodeficiency virus (HIV), human T-lymphotropic virus (HTLV), hepatitis A virus, hepatitis B virus, hepatitis C virus, hepatitis E virus, cytomegalovirus (CMV), Epstein-Barr virus (EBV), rubella virus, or treponema pallidum.

**FMT Capsule Preparation**

Fresh fecal samples were collected into a sterile container, sealed hermetically, and transferred/transported immediately to the manufacturing facility within two hours. Approximately 100g of feces was processed separately for each donor without pooling. All subsequent processes were conducted under anaerobic conditions. Feces were diluted and homogenized with sterile normal saline. The mixture was filtered and then centrifuged, and the pellet was resuspended with residual liquid and sterile proprietary live-cell protection solution. The suspension was then transferred into capsule shells. After the capsules (#0) were closed, a second encapsulation (#00) was followed. Therefore, each final FMT capsule had two layers of shells. The FMT capsules were frozen and stored below -75°C prior to being shipped to the research center on dry ice with a continuous temperature monitoring d evice. Upon arrival, the FMT capsules were immediately stored in a freezer that is below -75°C.

**Microbiome Analysis**

DNA extraction was performed using the DNeasy PowerSoil Pro Kit (QIAGEN, Shenzhen). Metagenomics libraries were sequenced for 150bp paired end reads using Illumina Novaseq6000.

When analyzing strain level colonization with Strainphlan3, using the distance matrix and the associated metadata, a threshold defining identical strains was inferred by selecting the top 1% from the distribution of the non-related-samples distances. The strain engraftment was inferred with the following rules: a strain from the donor was defined as "gain" when it presented in the recipient only after FMT treatment; When a strain from the donor was detected in the recipient after FMT treatment, and another strain within the same species existed in baseline samples, the strain was defined as "replace".

In terms of genome level colonization, species DNA sequence dissimilarity for a pair of samples was calculated, and the transmission of bacterial strains from donor to patient was calculated.

**Peripheral Blood TCR Sequencing Method**

RNA extraction on peripheral blood was conducted using the Tempus™ Blood RNA Tube and Tempus™Spin RNA Isolation Kit (Thermo Fisher). The extracted RNA was reverse transcribed into cDNA using the SuperScript™ IV VILOTM Master Mix (Thermo Fisher). Target gene amplification was conducted using Oncomine™ TCR Beta-SR Panel and Ion AmpliSeq™ Library Kit Plus (Thermo Fisher). Following digestion, junction identification, library amplification, and quantification of the final library product was performed using qPCR; the Ion S5 sequencing platform (Life) was used for TCR sequencing. Sequencing data was uploaded to Ion Reporter, an online platform for the data analysis. Subsequently, the alignment of TCR sequence and the comparison between the samples was performed.

**Peripheral Blood Flow Cytometry Sequencing Method**

Fresh peripheral blood samples were collected from patients at scheduled visits and transported to a Contract Research Organization (CRO) at 2~8°C for flow cytometric analysis. In panel 1 staining, the collected blood was activated with Leukocyte Activation Cocktail for 4 hours before being lysed and labeled with Fixable Viability Stain 450 to assess cell viability. After washing, the cells were stained with fluorescence dye-conjugated antibodies against surface markers (CD3, CD4, CD8, CD279,

Biolegend or BD Biosciences) for 15-20 minutes. Following fixation and permeabilization, cells were incubated with intracellular markers (Foxp3, Ki67, IFN-γ, BD Biosciences) for 15-20 minutes.

In panel 2, the peripheral blood was lysed and stained with 6 antibodies against surface markers (CD3, CD4, CD8, CD56, CCR7, CD45RA, BD Biosciences). The cells were then fixed, permeabilized, and stained with the intracellular marker CD68 (BD Biosciences). After washing, flow cytometry data was acquired using the NovoCyte 3130 instrument (ACEA Biosciences) and data analysis was performed using FlowJo software v.10.8.1 (BD Biosciences).

For comparing the flow cytometry data, baseline sample of LH001 was removed from analysis due to extremely small number of cells were detected.

**Validation Cohort Patients and Microbiome Analysis of Fecal Samples**

The validation cohort was based on an observational cohort focusing on gut microbiome profiling of GI cancer patients on immunotherapy who were hospitalized and scheduled for immunotherapy in Peking University Cancer Hospital since March 2018. This study was approved by the Ethics Committee at Peking University Cancer Hospital and all patients enrolled signed the informed consent. All tests and procedures were conducted in accordance with the Declaration of Helsinki. Patients were assigned to one of the following ICI treatment regimens without antibiotic application during the treatment, and until disease progression or intolerable toxicity: 1) PD-1/PD-L1 inhibitor, repeated every 2 or 3 weeks; 2) Combined PD-1/PD-L1 inhibitor and CTLA-4 inhibitor immunotherapy, repeated every 3 or 6 weeks. A total of 1443 fecal samples from 570 patients were profiled for gut microbiome composition using metagenomics sequencing. For validation, only gastric cancer patients with immunotherapy were selected, resulting in a total of 63 fecal samples from 63 patients.

**Method to Find Differential abundance bacteria between Rs and NRs**

Bacteria related to ICI treatment outcome were investigated from three ways. The first way only considered prevalence and abundance. In each sample, the relative abundance of bacteria was sorted and the ones added up to 80% were considered as the core microbiome of that sample. For each patient, the core microbiome existed in more than 60% of his/her samples were regarded as the core microbiome of that patient. The union of the core microbiome of the patient from the same treatment group were taken as the core microbiome of that group (NR, PRR, R). For the bacteria which were the core microbiome in both NR and R groups, the mean relative abundance of that bacteria in each group were calculated and the bacteria was added to the group in which its average relative abundance was higher. The second way performed a differential abundance analysis using the samples collecting in the mid-term evaluation timepoint, as all patients had their samples collected in this timepoint and got their respective clinical evaluation outcome. The third way included samples collected in a longer timeframe (baseline to 10 weeks after FMT treatment) and a mixed effect model was applied with patient added as a random effect. Time factor was limited to within 10 weeks after treatment to avoid imbalance in sampling time between different treatment groups (mean of sample collecting weeks after treatment: NR = 9.75, R = 23.5).
